# The contribution of Neanderthal ancestry to cancer risk

**DOI:** 10.64898/2026.11.10.26367742

**Authors:** Colin M. Brand, Yaen Chen, John A. Capra

## Abstract

Modern human genomes harbor DNA inherited from interbreeding with Neanderthals. Genetic variants inherited from Neanderthals associate with variation in many immune and metabolic phenotypes, two systems with direct relevance to cancer. However, the influence of Neanderthal variants on germline cancer risk has not been comprehensively evaluated.

Here, we evaluated genome-wide association study (GWAS) hits for 17 cancer types for Neanderthal ancestry. Twelve cancers have multiple Neanderthal alleles significantly associated with variation in risk. Next, we fine-mapped these risk loci and identified 32 associations across seven cancers where Neanderthal variants are likely causal (*≥* 50% posterior inclusion probability). Fine-mapped loci include Neanderthal alleles that both increase and decrease cancer risk. Nine of the 32 Neanderthal associations, including the high frequency *BNC2* Neanderthal variants, have signatures of adaptive introgression, and several suggest evidence of both mediated and antagonistic pleiotropy. Finally, we tested for enrichment of cancer heritability in genomic regions with Neanderthal ancestry. As expected, Neanderthal variants are depleted of heritability for most cancers. However, Neanderthal variants were enriched for heritability in breast cancer GWAS in both East Asian and Europeans, and this enrichment replicated using individual-level UK Biobank data.

These results suggest that Neanderthal alleles that influence germline cancer risk were largely selected against. However, introgressed Neanderthal variants implicated in many cancers persist, potentially due to adaptive pleiotropic effects. This evolutionary perspective provides new insights into the complex evolution and etiology of cancer risk among modern human populations.

## INTRODUCTION

The genomes of modern humans reflect complex demography and migration patterns over the past hundreds of thousands of years, including interbreeding with other hominin lineages, such as Neanderthals (Green et al., 2010; Nielsen et al., 2017). For example, following the most recent large human migration out of Africa, modern humans interbred with Neanderthals approximately 47,000 years ago (Fig. 1A, Iasi et al., 2026). As a result, DNA of Neanderthal ancestry comprises roughly 1-3% of most modern non-African genomes (Prüfer et al., 2014). However, the distribution of this Neanderthal ancestry is highly variable between and within populations. The total remaining Neanderthal ancestry across modern humans covers *≥*60% of the Neanderthal genome, revealing a complex landscape of selection following interbreeding (Harris and Nielsen, 2016; Petr et al., 2019).

**Fig. 1.**
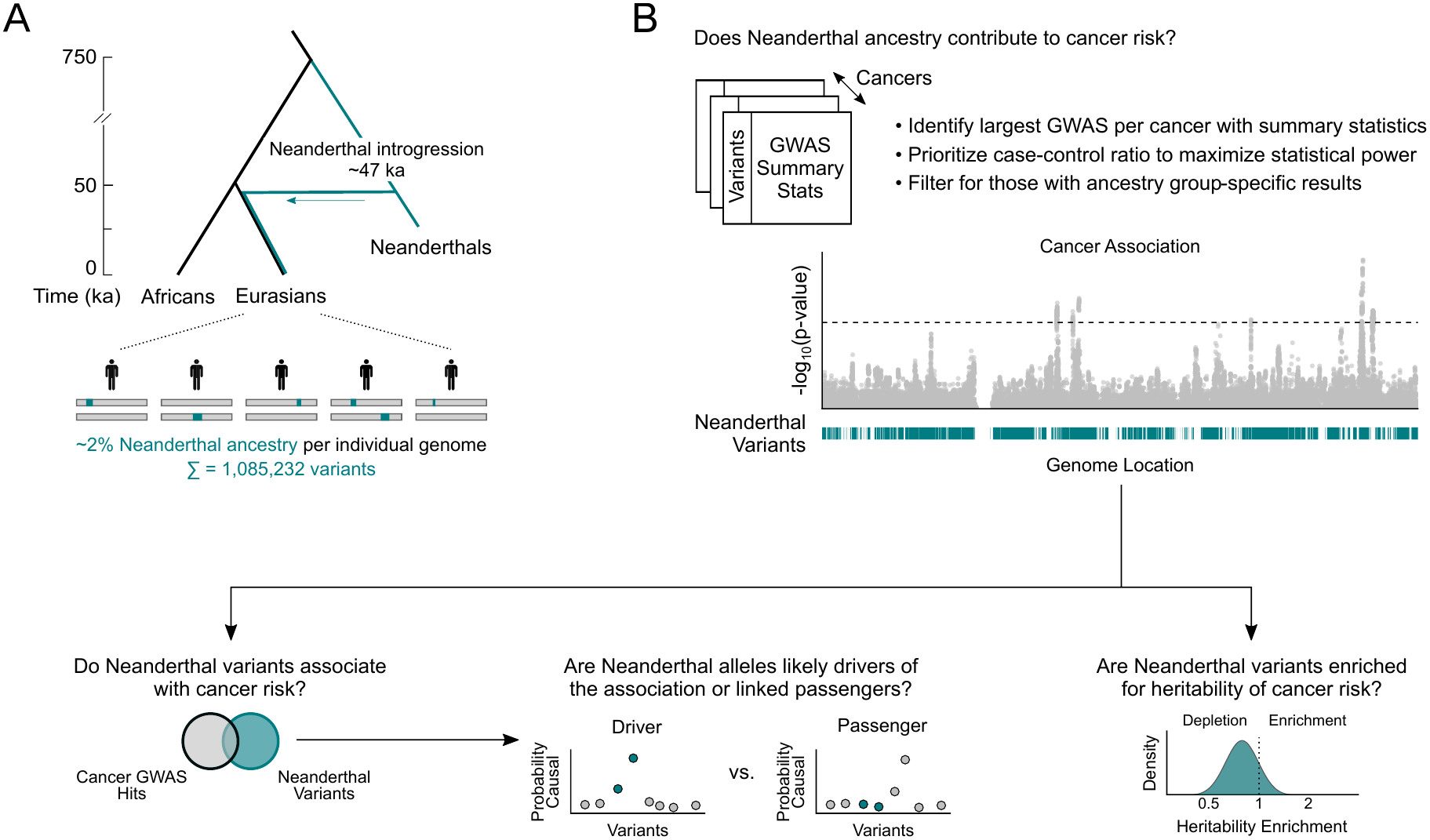
Strategy for quantifying the contribution of Neanderthal variants to cancer risk. (A) Due to interbreeding 47,000 years ago, Neanderthal DNA comprises 2% of the genome of most non-African individuals. As a result, more than one million (N = 1,085,232) variants of Neanderthal ancestry are present in modern human populations. (B) Evaluating introgressed Neanderthal variants for associations with germline cancer risk. Because Neanderthal ancestry is largely restricted to non-African populations, we curated 29 large cancer GWAS in the two ancestry groups with sufficiently powered studies, East Asian and European (Table 1). We then intersected these GWAS with maps of Neanderthal variants to identify associations, conducted fine-mapping on associated loci to identify putatively causal variants, and ran stratified LD-score regression to estimate their contribution to cancer heritability.

**Table 1.** Summary of cancer GWAS analyzed. Overview of the 29 cancer GWAS analyzed, including cancer type, study, ancestry group, and the number of cases, controls, and and total sample size. EAS = East Asian, EUR = European.

| **Cancer** | **Study** | **Ancestry Group** | **N Cases** | **N Controls** | **Total N** |
|---|---|---|---|---|---|
| Basal Cell Carcinoma | Seviiri et al. 2022 | EUR | 20,791 | 286,893 | 307,684 |
| Bladder Cancer | Rashkin et al. 2020 | EUR | 2,242 | 410,350 | 412,592 |
| Breast Cancer | Jia et al. 2025 | EAS | 21,319 | 106,116 | 127,435 |
| Breast Cancer | Zhang et al. 2020 | EUR | 133,384 | 113,789 | 247,173 |
| Colorectal Cancer | Lu et al. 2019 | EAS | 23,572 | 48,700 | 72,272 |
| Colorectal Cancer | Fernandez-Rozadilla et al. 2022 | EUR | 78,473 | 107,143 | 185,616 |
| Endometrial Cancer | Sakaue et al. 2021 | EAS | 1,200 | 60,614 | 61,814 |
| Endometrial Cancer | O’Mara et al. 2018 | EUR | 12,906 | 108,979 | 121,885 |
| Esophageal Cancer | Sakaue et al. 2021 | EAS | 1,388 | 159,201 | 160,589 |
| Esophageal Cancer | Rashkin et al. 2020 | EUR | 1,091 | 410,350 | 411,441 |
| Gastric Cancer | Sakaue et al. 2021 | EAS | 7,921 | 159,201 | 167,122 |
| Gastric Cancer | Zhou et al. 2018 | EUR | 554 | 393,372 | 393,926 |
| Liver Cancer | Sakaue et al. 2021 | EAS | 418 | 159,201 | 159,619 |
| Liver Cancer | Zhou et al. 2018 | EUR | 344 | 393,372 | 393,716 |
| Lung Cancer | Sakaue et al. 2021 | EAS | 4,444 | 159,201 | 163,645 |
| Lung Cancer | McKay et al. 2017 | EUR | 29,266 | 56,450 | 85,716 |
| Melanoma | Rashkin et al. 2020 | EUR | 6,777 | 410,350 | 417,127 |
| Ovarian Cancer | Sakaue et al. 2021 | EAS | 843 | 60,614 | 61,457 |
| Ovarian Cancer | Rashkin et al. 2020 | EUR | 1,259 | 410,350 | 411,609 |
| Pancreatic Cancer | Sakaue et al. 2021 | EAS | 499 | 159,201 | 159,700 |
| Pancreatic Cancer | Rashkin et al. 2020 | EUR | 663 | 410,350 | 411,013 |
| Prostate Cancer | Wang et al. 2023 | EAS | 10,809 | 95,790 | 106,599 |
| Prostate Cancer | Wang et al. 2023 | EUR | 122,188 | 604,640 | 726,828 |
| Renal Cancer | Purdue et al. 2024 | EAS | 621 | 86,796 | 87,417 |
| Renal Cancer | Purdue et al. 2024 | EUR | 25,890 | 743,585 | 769,475 |
| Squamous Cell Carcinoma | Seviiri et al. 2022 | EUR | 7,402 | 286,892 | 294,294 |
| Testicular Cancer | Zhou et al. 2018 | EUR | 2,981 | 401,788 | 404,769 |
| Thyroid Cancer | Sakaue et al. 2021 | EAS | 361 | 178,362 | 178,723 |
| Thyroid Cancer | Zhou et al. 2022 | EUR | 6,015 | 1,333,754 | 1,339,769 |

Although Neanderthal DNA remains largely in genomic regions under low evolutionarily constraint, Neanderthal alleles are associated with many traits in various biological domains (Dannemann and Kelso, 2017; McArthur, Rinker, and Capra, 2021; Simonti et al., 2016; Wei et al., 2023). These traits include many immune and metabolic phenotypes (Abi-Rached et al., 2011; Dannemann, Andrés, and Kelso, 2016; Gittelman et al., 2016; Jagoda et al., 2022; Mendez, Watkins, and Hammer, 2012; Quach et al., 2016; Sams et al., 2016; Williams et al., 2014; Zeberg and Pääbo, 2020; Zeberg and Pääbo, 2021; Zhou et al., 2021), two systems with direct relevance to cancer risk and progression (Andrejeva and Rathmell, 2017; Moretti, Cioccoloni, and Perucha, 2025). Neanderthal ancestry has been proposed to promote somatic mutations in liver cancer (Taravella Oill, Buetow, and Wilson, 2022), but the contribution of Neanderthal variants to germline cancer risk has not been comprehensively evaluated.

Here, we quantify the contribution of Neanderthal variants to cancer risk by integrating GWAS for 17 cancers with Neanderthal ancestry maps. First, we identify cancers whose significantly associated loci harbor Neanderthal alleles. Second, we conduct fine-mapping to nominate putatively causal Neanderthal variants. Third, we highlight multiple fine-mapped example variants, including Neanderthal alleles that influence breast cancer and skin cancer risk but have positively selected pleiotropic effects. Finally, we quantify enrichment for heritability of cancer risk among regions of Neanderthal ancestry using summary statistics and replicate our results using individual-level genotype data.

## RESULTS

### Characterizing germline cancer risk in the context of Neanderthal ancestry

We identified autosomal Neanderthal ancestry variants based on four previous maps of Neanderthal introgression with single-nucleotide variant (SNV) resolution: Browning et al. 2018, Sankararaman et al. 2016, Skov 2024, and Vernot et al. 2016. Analyzing multiple Neanderthal variant sets improves the robustness of downstream conclusions (Chen, Velazquez-Arcelay, and Capra, 2026). The resulting maps comprise 1,085,232 variants in total, with individual maps contributing between 232,262 and 892,930 variants after filtering (Fig. S1). While the frequency of many Neanderthal variants is relatively low in modern human genomes, between 333,402 (East Asian) and 438,325 (South Asian) of the 912,978 called variants with available frequency estimates—37% to 48%—have a minor allele frequency *≥* 1% within a given non-African gnomAD ancestry group (Fig. S2). Hundreds of thousands of these variants are therefore common enough to be tested for phenotype associations in GWAS.

We identified germline risk variants for cancer types using publicly available GWAS summary statistics (Table S1). We prioritized GWAS with a large discovery sample size and a high case-control ratio (Fig. 1B, Table 1). We also focused on GWAS that analyzed a single non-African ancestry group due to the challenges of interpreting multi-ancestry GWAS and the potential differences in Neanderthal ancestry patterns between ancestries. This resulted in 29 total cancer GWAS across 17 cancer types among individuals with genetic similarity to East Asians (N = 12) and Europeans (N = 17) (Table 1, Table S2).

### Loci associated with germline cancer risk harbor Neanderthal variants

We clustered significant cancer GWAS associations into independent risk loci based on linkage disequilibrium (LD) calculated from ancestry-specific *r*^2^ data from the 1000 Genomes Project (Auton et al., 2015). We resolved loci for the 20 GWAS with multiple genome-wide significant variants (*P* = 5 *×* 10^−8^) (Table S2). Next, we intersected the Neanderthal variants with the cancer GWAS risk loci and classified loci as potentially introgressed if they contain at least two Neanderthal variants from the union of all four introgression datasets (N = 1,085,232) (Fig. 2A).

**Fig. 2.**
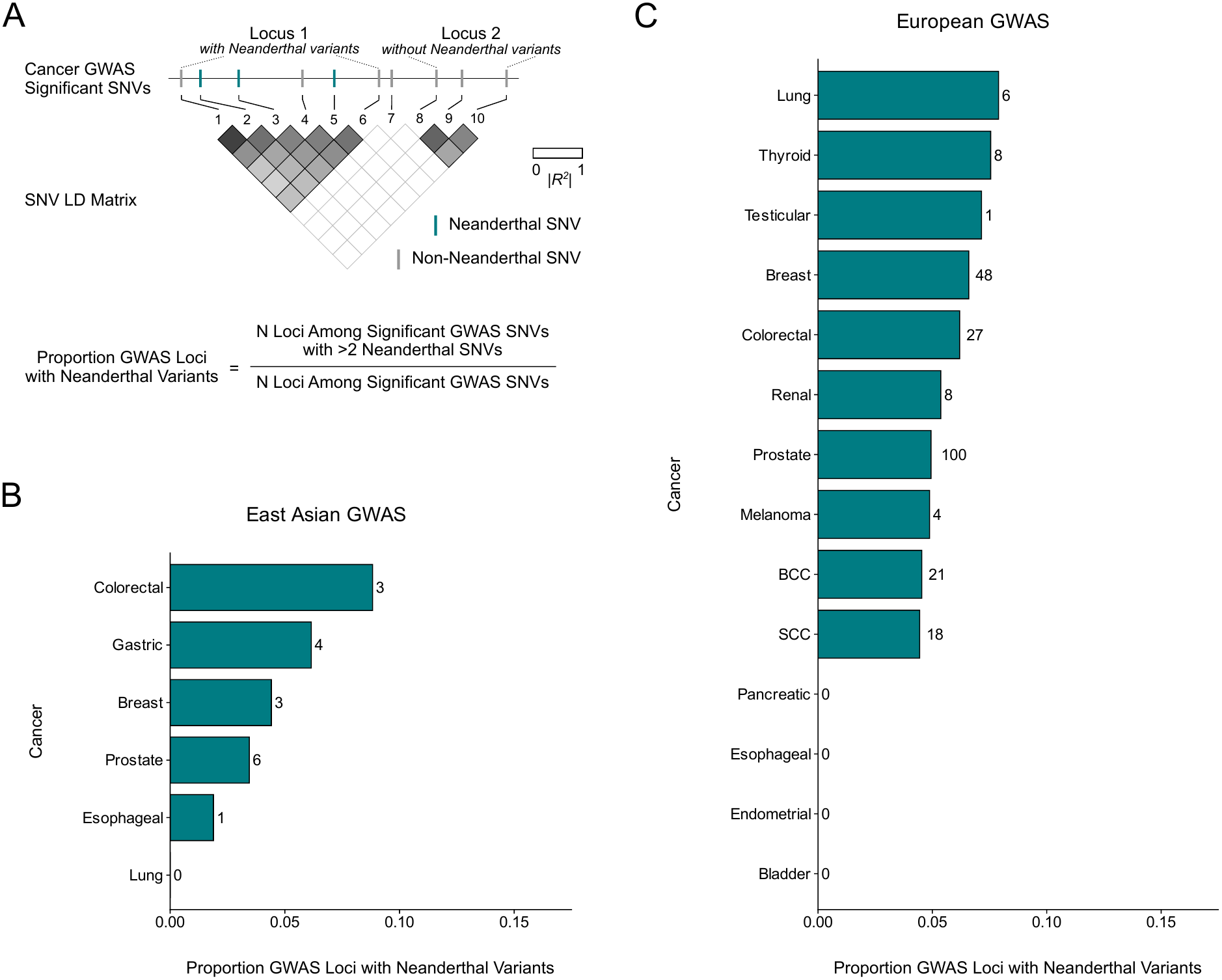
Cancer risk GWAS loci harbor Neanderthal variants. (A) Schematic of the approach for identifying candidate loci with Neanderthal variants. Cancer-associated loci were defined within each GWAS by LD clumping of genome-wide significant variants (*P* < 5 10^−8^) within 250-kb windows, using ancestry-matched reference panels from 1000 Genomes. Loci were required to comprise at least two variants at the given *r*^2^ threshold, since an association based on a single variant offers no secondary associations to assess. The same requirement was applied to Neanderthal variants, so that a locus is counted as harboring Neanderthal variants only when at least two Neanderthal variants are present; this reduces the influence of spurrious map calls and associations. (B) Five of six cancer GWAS among East Asian individuals have at least one significantly associated locus containing Neanderthal variants. N gives the number of such loci per GWAS. (C) 10 of 14 cancer GWAS among European individuals have at least one significantly associated locus containing Neanderthal variants. N gives the number of such loci per GWAS.

Overall, 15 out of 20 cancer GWAS contained at least one risk locus with Neanderthal variants (Fig. 2, Table S3). Stratifying by genetic ancestry group, loci with Neanderthal variants are present among five out of six cancer GWAS among individuals genetically similar to East Asians (Fig. 2B, Table S3) and 10 out of 14 cancer GWAS conducted among individuals genetically similar to Europeans (Fig. 2C, Table S3).

As expected, the number of potential Neanderthal associations is sensitive to the LD threshold used to define a locus (Fig. S3, Table S3). Many GWAS typically use a permissive LD threshold, such as *r*^2^ > 0.2. This approach yields higher proportions of loci with Neanderthal variants but fewer loci overall. A stricter LD threshold of *r*^2^ = 0.8 results in lower proportions of loci with Neanderthal variants, but more cancer-associated loci overall. Therefore, we chose *r*^2^ = 0.5 for downstream analyses to balance the tradeoff between loci containing Neanderthal variants and overall number of independent loci.

### Fine-mapping nominates Neanderthal alleles as causal cancer-risk variants

The co-localization of germline cancer risk variants and Neanderthal variants could reflect causal roles for the Neanderthal alleles. However, it is also possible that the Neanderthal variants are simply in high LD with causal variants that mutated onto Neanderthal haplotypes after introgression (Rinker et al., 2020; Skov et al., 2020). To resolve these scenarios, we used statistical fine-mapping to estimate whether Neanderthal variants are among the credible sets for causally influencing germline cancer risk.

First, we identified genome-wide significant loci in each GWAS with Neanderthal variants (Fig. 3A). Within each GWAS, we started with the most strongly associated Neanderthal variant, included all Neanderthal variants within 500 kb, and then moved on to the next most strongly associated remaining variant to define the next locus, and so on. We then fine-mapped the *±*500 kb window around each index variant with SuSiE-RSS, using ancestry-matched 1000 Genomes LD reference panels. We fine-mapped 165 loci, of which 152 converged or were rescued by refitting (Table S4). These comprised 448 credible sets across 148 loci, 306 of which contained a variant reaching genome-wide significance (|*Z*| *≥* 5.45, *P* < 5 *×* 10^−8^) (Table S5). Collapsing sets based on overlapping windows left 302 distinct associations, 70 of which contained at least one Neanderthal variant (Table S5).

**Fig. 3.**
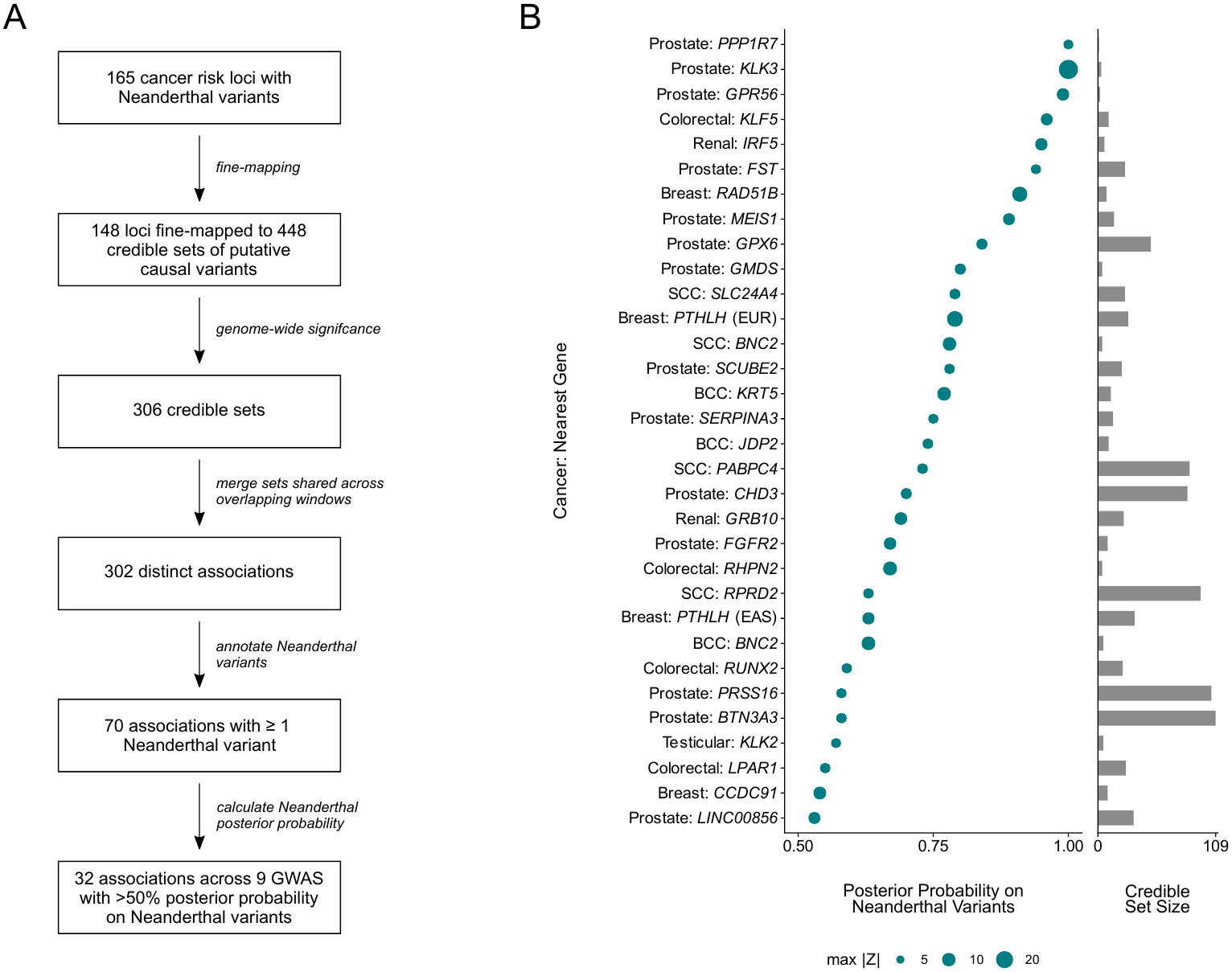
Fine-mapping reveals that Neanderthal variants drive associations with cancer risk at many loci. (A) Workflow for fine-mapping cancer risk loci with significant Neanderthal GWAS variants. (B) 32 associations across 9 GWAS for which the posterior probability that a Neanderthal variant is causal exceeds 50%. Point size indicates the maximum association strength (*Z*-score) among the Neanderthal variants in the credible set; bars indicate the number of variants in the credible set. Loci are identified by the nearest gene. BCC = basal cell carcinoma, SCC = squamous cell carcinoma, EAS = East Asian, EUR = European.

In 32 of these 70 associations, the posterior probability that the causal variant is a Neanderthal variant exceeded 50% (Fig. 3B, Tables S6-7). The 32 associations span nine GWAS and seven cancers: basal cell carcinoma, breast, colorectal, prostate, renal, squamous cell carcinoma, and testicular cancer. Previous work has demonstrated that introgressed Neanderthal variants are associated with and enriched for effects on integumentary and pigmentation phenotypes (Vernot and Akey, 2014; Racimo et al., 2015; Simonti et al., 2016; Dannemann and Kelso, 2017; McArthur, Rinker, and Capra, 2021). Thus, we were particularly curious whether Neanderthal variants contribute to the risk landscape for skin cancers. Indeed, seven of the 32 Neanderthal-majority associations, across six loci, are in skin cancers: two unique to basal cell carcinoma (BCC), three unique to squamous cell carcinoma (SCC), and an association in the *BNC2* locus, which is shared by both (Fig. 3B). However, we note that these associations are also influenced by GWAS discovery power; better-powered studies are more likely to have Neanderthal variant associations.

Two Neanderthal variant sets associate with multiple cancers: the *KLK* cluster on 19q13.33 associates with both prostate and testicular cancer, and the *BNC2* locus with basal and squamous cell carcinoma. In both cases, the contributing studies share cohorts, so neither is a fully independent observation. Moreover, the prostate cancer associations at *KLK* are potentially influenced by prostate-specific antigen (PSA) ascertainment bias (Kachuri et al., 2023). The *PTHLH* association is the only one recovered in both ancestry groups, from two breast cancer GWAS of unrelated samples, one of individuals with genetic similarity to Europeans and one to East Asians.

### Some Neanderthal variants increase cancer risk, while others decrease cancer risk

Of the 32 high-confidence Neanderthal cancer associations, 14 increase risk and 18 decrease risk. For some cancers, different Neanderthal associations have opposing effects. For example, in SCC, Neanderthal variants in the gene *BNC2* increase risk (*β* = 0.063). This gene encodes a zinc-finger transcription factor that is involved in many cellular processes and influences skin pigmentation (Jacobs et al., 2013). In contrast, Neanderthal alleles at *SLC24A4* decrease risk for SCC (*β ≈* –0.055; Fig. S4, Table S7). The *SLC24A4* Neanderthal variants also overlap a known pigmentation locus (Han et al., 2008); however, they do not overlap pigmentation credible sets (Table S8). Instead, the Neanderthal variants (e.g., rs79342051) are associated with increased *SLC24A4* expression in skeletal muscle (Fig. S5) and with higher IGF-1 levels and lower smoking initiation (Table S8). Thus, while both sets of Neanderthal variants influence genes involved in skin pigmentation, their effects on SCC risk are in opposite directions.

### Many causal cancer-associated Neanderthal loci have evidence of adaptive introgression

Five of the 32 Neanderthal cancer associations intersect regions previously identified as adaptively introgressed: variants near *PABPC4* for squamous cell carcinoma, *BNC2* for both basal and squamous cell carcinoma, the type II keratin cluster on chromosome 12 for basal cell carcinoma, and variants within *LPAR1* for colorectal cancer (Table S9). Four more fall within 30 kb of a region with evidence of adaptive introgression: variants within *RHPN2* for colorectal cancer, within *RAD51B* for breast cancer, and near *PTHLH* for breast cancer in both ancestry groups (Table S9). We examine two of the cancer-associated loci with evidence of adaptive introgression (*BNC2*, *PTHLH*) in more detail in the following sections.

Notably, four of the five cancer-associated loci overlapping evidence of adaptive introgression function in the hair and skin. High frequency Neanderthal variants in the *BNC2* locus that are implicated in many studies of adaptive introgression associate with increased risk for both BCC and SCC (see below). In addition, adaptively introgressed Neanderthal alleles associate with decreased risk (rs687751 *β* = –0.100) for BCC in the type II keratin cluster on chromosome 12. Meanwhile, Neanderthal variants near *PABPC4* on chromosome 1 associate with increased SCC risk (rs3768321 *β* = 0.041) (Fig. S6). The overall enrichment for Neanderthal variant contributions to hair and skin phenotypes has been proposed to be the result of adaptive benefits after introgression (McArthur, Rinker, and Capra, 2021). This suggests that adaptive benefits potentially due to pleiotropic effects may have contributed to the maintenance of these cancer-associated Neanderthal variants.

### Adaptively introgressed Neanderthal variants in *BNC2* associate with increased skin cancer risk, consistent with mediated pleiotropy

The *BNC2* locus harbors a Neanderthal haplotype occurring at high frequency among Europeans. This region has been consistently identified as adaptively introgressed in Europeans (Racimo et al., 2015; Vernot and Akey, 2014; Gower et al., 2021; Zhang et al., 2023; Iasi et al., 2026). The haplotype is largely absent among East Asians, despite East Asians carrying the highest average Neanderthal ancestry among continental groups (Vernot and Akey, 2014).

We identified seven credible sets near *BNC2* for BCC and two for SCC. The strongest association in both GWAS is a six-variant set spanning 16.86–16.88 Mb that contains no Neanderthal variants (|*Z*| up to 14.0). The strongest Neanderthal association signal comes from a separate credible set 60 kb upstream (Fig. 4A) that is present in both BCC and SCC GWAS and tagged by the same lead variant (rs2026805). The Neanderthal allele at rs2026805 associates with increased cancer risk in both GWAS (*β* = 0.104 in BCC and 0.063 in SCC). Five variants are associated in BCC, and a subset of four are found in SCC. The posterior probability that the causal variant is Neanderthal is 0.63 for BCC and 0.78 for SCC (Figs. 4B, S7).

**Fig. 4.**
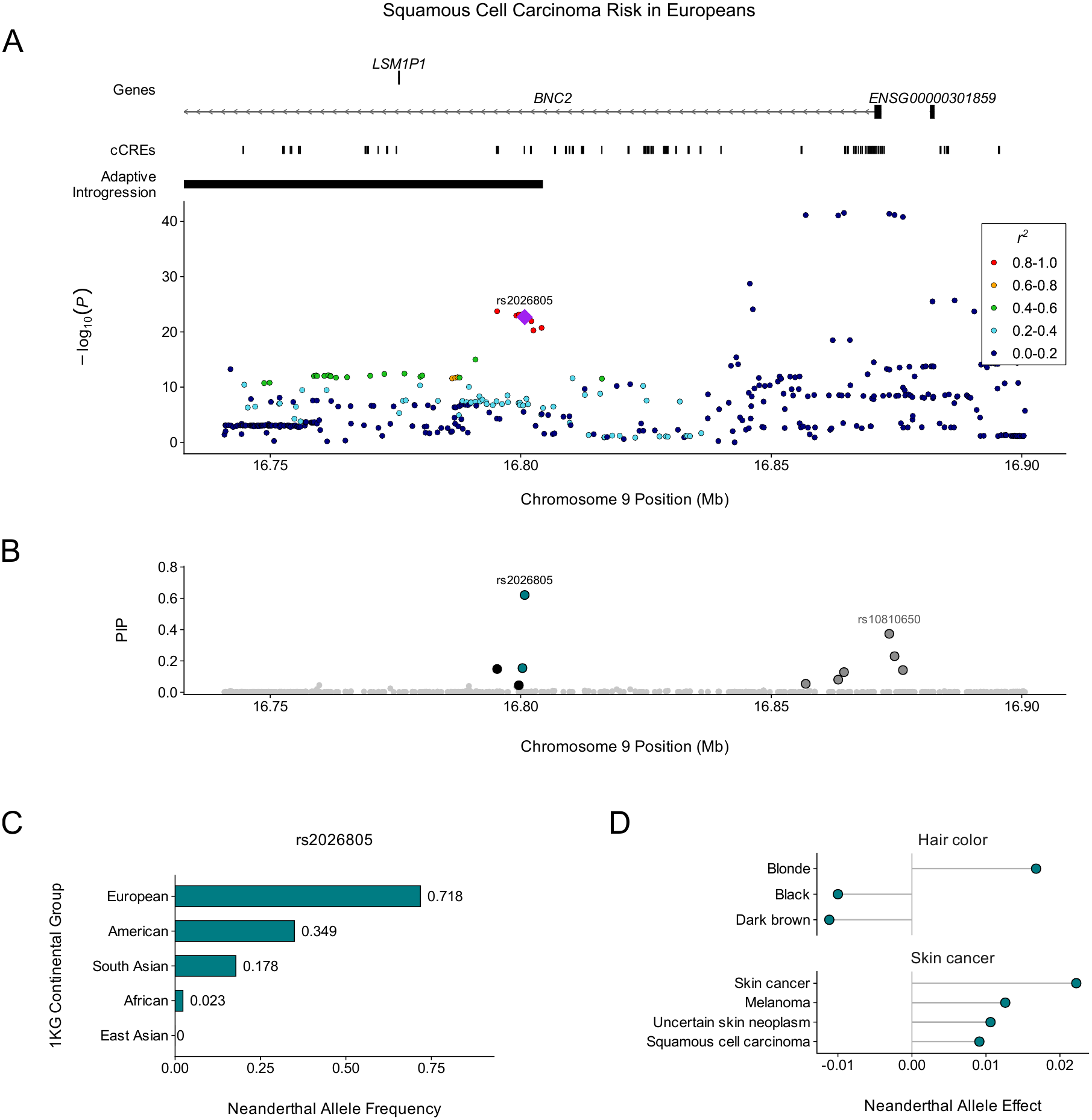
Adaptively introgressed Neanderthal variants within *BNC2* associate with increased skin cancer risk. (A) LocusZoom plot of variants from the Seviiri et al. (2022) squamous cell carcinoma GWAS conducted among individuals with genetic similarity to Europeans in 1000 Genomes. All variants in the summary statistics within the window are shown. Binned LD (*r*^2^) with rs2026805 is indicated by color; variants absent from the 1000 Genomes reference panel, for which *r*^2^ could not be calculated, are plotted in the lowest bin. The index variant, rs2026805, is indicated by a purple diamond. cCREs, as profiled by ENCODE, and adaptively introgressed regions from Iasi et al. (2025) are shown above. (B) Posterior inclusion probabilities (PIPs) for the variants in A. Teal and black mark members of CS2, the credible set this figure concerns, split by whether the variant is Neanderthal (teal) or not (black). Dark grey marks members of the other credible set at the locus (CS1), and light grey variants belong to no credible set. (C) Neanderthal allele frequencies for rs2026805 among 1000 Genomes samples, stratified by continental group. (D) Effects of the Neanderthal haplotype tagged by rs2026805 on hair color and skin cancer traits in Open Targets. Each point is one study–trait pair from an Open Targets credible set containing rs2026805. Betas are those of each credible set’s lead variant, whose alternate allele lies on the Neanderthal haplotype (lead variant in our *BNC2* cancer credible sets or in high LD with rs2026805, r² ≥ 0.91 in 1000 Genomes Phase 3 EUR), so positive values indicate an increase associated with the Neanderthal allele. Hair color traits are binary categories (UK Biobank field 1747). Effect sizes are on study-specific scales and are not comparable across traits.

In both sets, the highest-PIP variants, rs2026805 and rs2026804, are from Neanderthals and overlap an adaptively introgressed interval (Figs. 4A, 4B, S7) (Iasi et al., 2026). The remaining members are not called by any Neanderthal map but segregate on the same haplotype at comparable frequencies: rs10962605 and rs10962599 in both sets, and rs10810636 in BCC (Fig. 4C). Thus, these variants may also be Neanderthal in origin, and the Neanderthal posterior probabilities at this locus are likely even higher. No common, frequency-compatible structural variant (SV) was identified in gnomAD-SV v4 within the locus (Table S10) nor in meaningful linkage disequilibrium with the Neanderthal credible set variants at *BNC2* (maximum *r*^2^ = 0.15 between any SV and rs2026805; Table S11).

Next, we analyzed functional data to interpret the consequences of the Neanderthal variants. Two of four SCC-associated variants, rs2026805 and rs10962599, intersect candidate cis-regulatory elements (cCREs), in particular distal enhancer-like elements, identified by ENCODE (Fig. 4A). While we did not identify any expression quantitative-trait loci (eQTL) or splicing quantitative trait loci (sQTL) in GTEx, the credible set variants for cancer are also in credible sets for pigmentation and other skin cancer traits (Fig. 4D, Table S12). Consistent with our observations, the Neanderthal allele associates with increased skin cancer risk, lighter hair color (Fig. 4D), and lighter skin color (Table S12).

Lighter skin pigmentation increases skin cancer risk, but among humans living at higher latitudes, it is an adaptation that balances photoprotection against the need for vitamin D synthesis (Jablonski and Chaplin, 2000). This adaptation was potentially enabled by Neanderthals, who occupied higher latitudes for hundreds of thousands of years before the most recent *Homo sapiens* emigrations from Africa. In ancient DNA, the haplotype is detected in European and West Asian specimens by *∼*13,700 years ago at frequencies near present-day levels (Fig. S8). Collectively, these patterns are consistent with a scenario of mediated pleiotropy, where an adaptively introgressed Neanderthal haplotype that resulted in lighter pigmentation also increases risk of skin cancers via this effect.

### A Neanderthal haplotype at *PTHLH* associates with reduced breast cancer risk in both East Asians and Europeans

Neanderthal variants account for more than half of the causal posterior probability at four breast cancer risk loci. For example, associations at *PTHLH* were discovered in two independent GWAS, one of individuals with genetic similarity to East Asians and one to Europeans. *PTHLH* encodes parathyroid hormone-related protein, which is produced by normal breast epithelium and causes hypercalcemia of malignancy, and the 12p11 region that contains it has been consistently identified as a breast cancer susceptibility locus (Ghoussaini et al., 2012; Zhang et al., 2020). Several other genes are present in the associated region, including *CCDC91* and a lncRNA (ENSG00000247934).

Fine-mapping resolved a single association in the East Asian GWAS: a 34-variant credible set spanning 28.140–28.170 Mb, of which 27 are Neanderthal variants; the posterior probability that the causal variant is Neanderthal is 0.633 (Figs. 5A, 5B). In the European GWAS, fine-mapping resolved three associations (Figs. 5A, 5C). Credible set 1 (CS1) is composed of a single strongly associated variant (rs7297051, |Z| = 17.34, PIP = 0.983) at 28.175 Mb and contains no Neanderthal variants. Credible set 2 (CS2) consists of nine variants spanning 28.318–28.447 Mb, including five Neanderthal variants. Credible set 3 (CS3) is a 28-variant credible set spanning 28.140–28.182 Mb, of which 22 are Neanderthal variants, including the locus index variant: rs788464 (Fig. 5C). The posterior probability that the causal variant is Neanderthal is 0.541 for CS2 and 0.789 for CS3. Twenty of the 28 European CS3 variants are also in the East Asian credible set, while eight are found only in the European GWAS and 14 are found only in the East Asian GWAS. Conditional analyses indicate three independent associations in the European GWAS and suggest a second, unresolved association in the East Asian GWAS (Fig. S9, Supplementary Text).

**Fig. 5.**
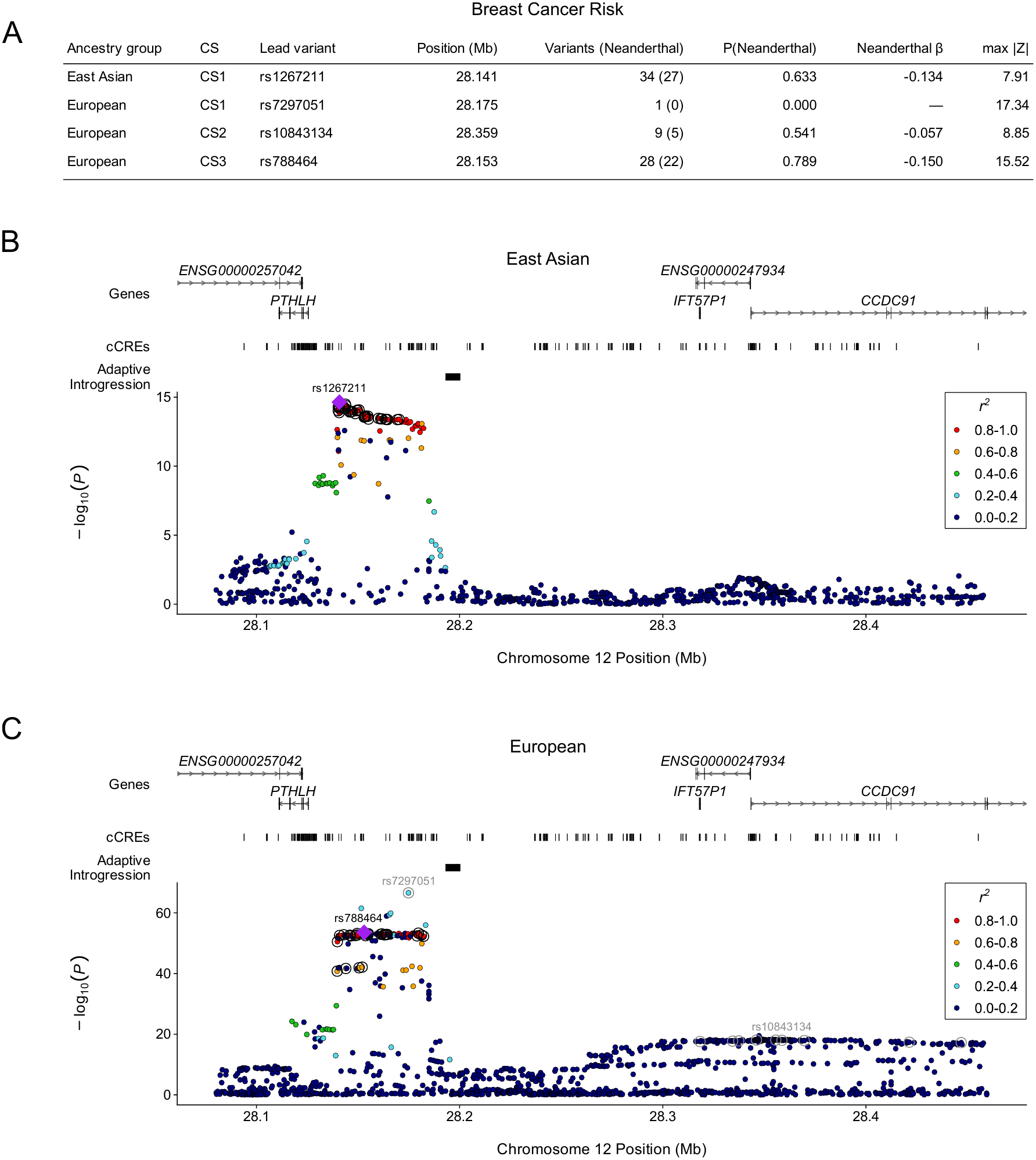
A Neanderthal haplotype at *PTHLH* associates with reduced breast cancer risk in both East Asians and Europeans. (A) Credible sets identified by SuSiE-RSS near *PTHLH* in both ancestry groups. Variants (Neanderthal) gives the number of variants in the credible set and, in parentheses, how many are Neanderthal. P(Neanderthal) is the posterior probability that the causal variant underlying the credible set is Neanderthal, summed over all Neanderthal variants in the fine-mapped window and so not restricted to the set’s members. Neanderthal *β* is the effect of the Neanderthal allele, taken from the highest-PIP member of the set carrying an allele call; it is undefined for European CS1, which contains no Neanderthal variants. (B) LocusZoom plot of variants from the Jia et al. (2025) breast cancer GWAS conducted among individuals with genetic similarity to East Asians in 1000 Genomes. All variants in the summary statistics within the window are shown. Binned LD (*r*^2^) with rs1267211 is indicated by color; variants absent from the 1000 Genomes reference panel, for which *r*^2^ could not be calculated, are plotted in the lowest bin. rs1267211, the lead variant of the East Asian credible set, is marked by a purple diamond, and the set’s remaining members are circled in black. This locus has no other credible set. Gene models are GENCODE, cCREs are from ENCODE, and adaptively introgressed regions are from Iasi et al. (2025). Gene models are limited to two rows, so some non-coding models in dense windows are not shown. (C) The same window in the Zhang et al. (2020) breast cancer GWAS conducted among individuals with genetic similarity to Europeans, visualized as in B with *r*^2^ relative to rs788464. rs788464, the lead variant of CS3, is marked by a purple diamond and the set’s remaining members are circled in black. Members of the locus’s other credible sets are circled in grey, with each set’s lead variant labelled in grey: rs7297051, the single member of CS1 and the most strongly associated variant in the region, and rs10843134, the lead of CS2. Both panels span the same coordinates so the two ancestry groups can be compared directly.

The Neanderthal alleles in these credible sets consistently associate with decreased risk of breast cancer (e.g., rs788464: East Asian *β* = –0.128 and European *β* = –0.150). Neanderthal variants in these credible sets are also eQTLs for *PTHLH* and the other nearby genes across several tissues, with effects in both directions depending on the gene and tissue (Supplementary Text, Fig. S10); which gene mediates risk at 12p11 remains unresolved. Nonetheless, our results link Neanderthal variants at this locus to altered expression of these genes and to reduced breast cancer risk. No common structural variant was in high linkage disequilibrium (highest observed *r*^2^ = 0.22) with the Neanderthal credible set variants at *PTHLH*; thus, structural variants that have been observed in this locus are not driving the association (Tables S13-14, Supplementary Text).

Several variants in the *PTHLH* credible sets with high PIP matched the Neanderthal allele and were in high LD with Neanderthal variants, but were not called as Neanderthal variants themselves because they were present on different haplotypes in African ancestry individuals (Fig. 6A). Previous work has shown that this pattern is characteristic of reintroduced ancestral alleles (RAAs) (Rinker et al., 2020). While most variants modern humans inherited from Neanderthals arose on the Neanderthal lineage, Neanderthal introgression also reintroduced hundreds of thousands of ancestral alleles that were lost during bottlenecks caused by emigration out of Africa (Fig. 6A). These RAAs are enriched for eQTLs and phenotype associations, and they also exhibit regulatory activity comparable to non-Neanderthal variants (Rinker et al., 2020). Intersecting the credible set variants with ancestry-group-specific RAAs, we discovered that 13 out of 34 East Asian variants are RAAs, whereas 7 out of 28 European CS3 variants are RAAs (Fig. 6B). rs788464, an RAA and the lead CS3 variant, segregates in African populations as well as outside them (Fig. 6C) and is an eQTL for *PTHLH* in skeletal muscle (Fig. 6D). Three further variants (rs1267211, rs1267213, rs788461), which include the highest-PIP member of the East Asian credible set, are identified as RAAs but not called by any introgression map, likely because many introgression detection methods miss RAAs due to their presence in African populations often used as outgroups. Thus, ancestral alleles reintroduced by introgression likely influence cancer risk at this locus and the posterior probabilities for Neanderthal contributions reported here are likely a lower bound.

**Fig. 6.**
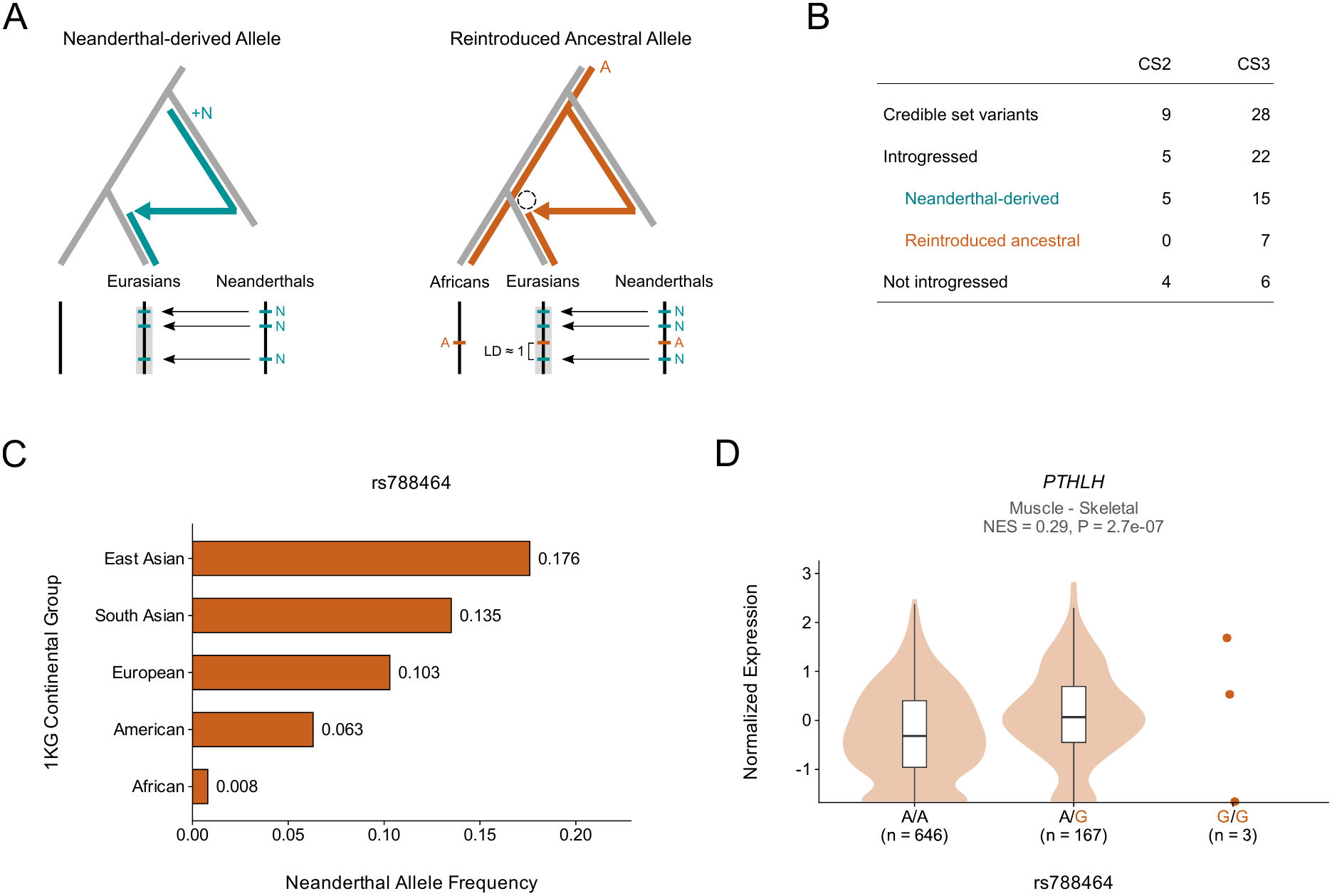
Reintroduced ancestral alleles are carried on the Neanderthal haplotype at *PTHLH* and associate with decreased breast cancer risk. (A) Many Neanderthal variants in modern humans are those that derived within the Neanderthal lineage (NDAs). However, Neanderthal interbreeding also reintroduced ancestral alleles lost during Out of Africa (RAAs). These alleles occur in modern Africans and are in perfect linkage disequilibrium with NDAs. (B) Summary of NDAs and RAAs among credible sets at *PTHLH*. The RAAs among CS3 include the index variant: rs788464. (C) Neanderthal allele frequencies for rs788464 among 1000 Genomes samples, stratified by continental group. (D) Normalized expression of *PTHLH* in skeletal muscle by rs788464 genotype in GTEx v10. Violins show the distribution of per-sample expression for each genotype, and boxes the median and interquartile range; the G/G genotype, with three samples, is shown as individual points. Sample sizes are given below each genotype. The reintroduced ancestral allele (G) is shown in orange. NES, normalized effect size of the G allele.

### Neanderthal variants are consistently enriched for breast cancer heritability in both East Asians and Europeans

Having identified Neanderthal variants within the fine-mapped credible sets for loci across multiple cancers, we next evaluated the overall contribution of Neanderthal ancestry to the heritability of each cancer compared to the expectation based on their frequency and genomic distribution. Neanderthal variants are largely depleted of heritability for most complex traits (McArthur, Rinker, and Capra, 2021; Wei et al., 2023); thus, we anticipated that they would contribute less than expected to most cancers. We quantified heritability enrichment for Neanderthal variants for each cancer GWAS using stratified LD score regression (S-LDSC) against the baseline model, using 1000 Genomes as the reference panel. We considered each of the four introgression maps separately for each GWAS to better evaluate robustness of the conclusions to the introgression mapping strategy used.

Of the 28 GWAS with available effect sizes, eight carried sufficient heritability signal for partitioned analysis (mean *χ*^2^ *≥* 1.02 and an observed-scale heritability *≥* 0.05): basal cell and squamous cell carcinoma, breast cancer in both ancestry groups, colorectal and lung cancer in Europeans, and prostate cancer in both ancestry groups (Fig. 7, Table S15). The remaining 20 were excluded for failure to meet one or both criteria. Thus, these results describe the best powered subset, rather than all cancers surveyed.

**Fig. 7.**
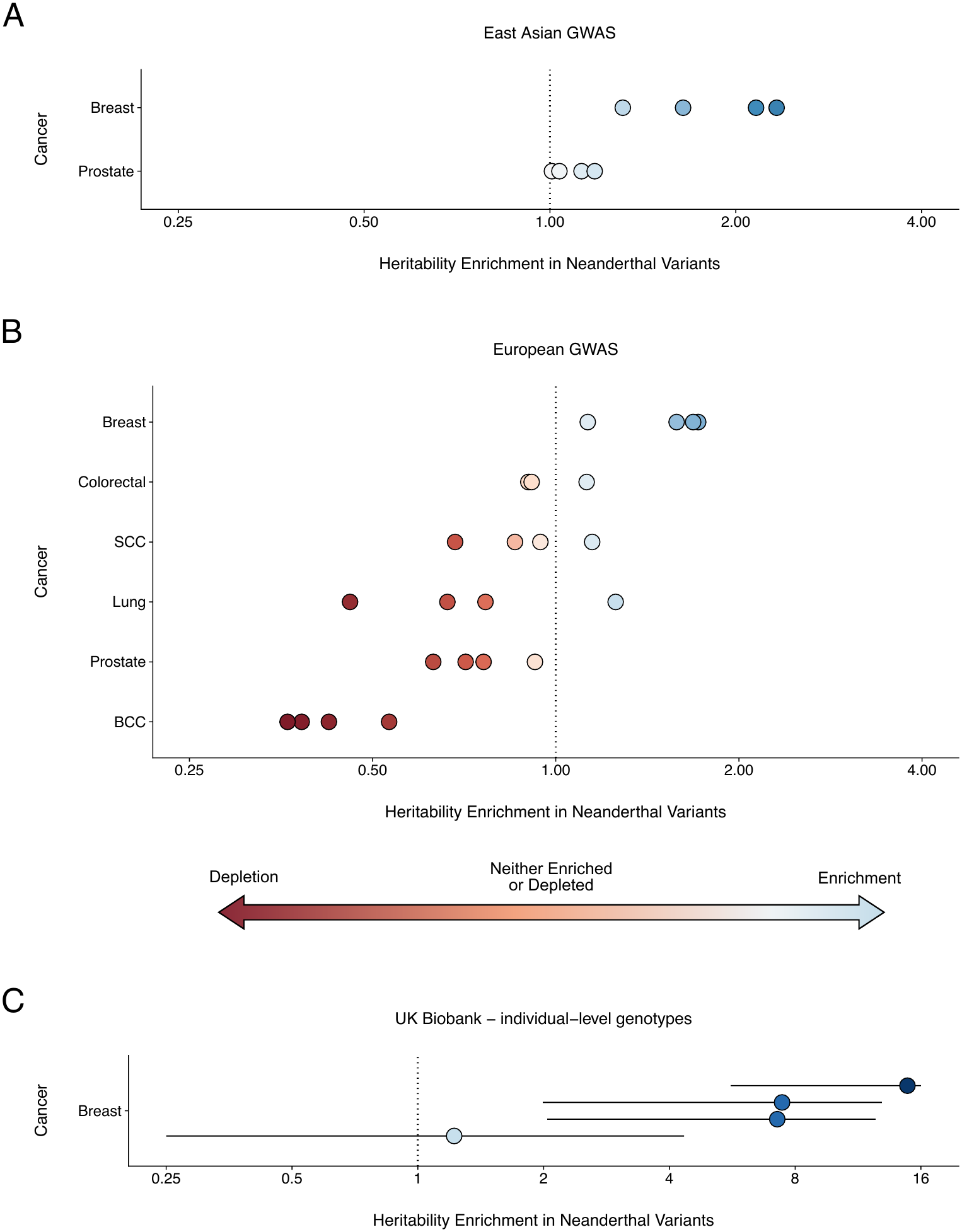
Neanderthal alleles are enriched for heritability of cancer risk in breast cancer. (A) Heritability enrichment calculated using ldsc for Neanderthal variants among East Asian cancer risk GWAS. Each point reflects enrichment for an individual introgression map: Browning et al., 2018, Sankararaman et al., 2016, Skov, 2024, and Vernot et al., 2016. We excluded any GWAS-introgression map pairs where mean *χ*^2^ < 1.02, estimated heritability was 0, and enrichment standard error was > 10. Color indicates heritability enrichment. The x-axis is log-scaled. Values < 0.125 are clipped for visualization. (B) Heritability enrichment calculated using ldsc for Neanderthal variants among European cancer risk GWAS. Data are visualized as in A. (C) Heritability enrichment for Neanderthal variants in breast cancer, estimated with GENIE from individual-level genotypes in a matched case-control cohort drawn from the UK Biobank. Bars indicate ±1 jackknife SE. Data are visualized as in A.

As expected, Neanderthal alleles were largely depleted or neutral in their contributions to the heritability of cancer risk (Table S16). Breast cancer was the exception: enrichment point estimates exceeded 1 for all four introgression maps in both East Asians (1.31–2.33; Fig. 7A) and Europeans (1.13–1.71; Fig. 7B), the only cancer for which this held in both ancestry groups. No individual estimate reached nominal significance (minimum *P* = 0.23 across the 32 tests among GWAS passing heritability QC; minimum Benjamini–Hochberg *q* = 0.95).

Given the breast cancer risk heritability enrichment among Neanderthal variants in published GWAS, we validated this signal using individual-level genotypes from the UK Biobank, which contributes to neither breast cancer GWAS analysed here. In a cohort of 14,346 cases and 40,126 matched controls, Neanderthal variants were enriched for breast cancer heritability across all four introgression maps, ranging from 1.22 to 14.85x, although no individual estimate was statistically significant (Pazokitoroudi et al., 2024; Fig. 7C).

These results suggest that Neanderthal variants are not enriched for contributions to risk of most cancers, but that they may contribute more than expected to breast cancer risk. Despite the relatively small number of loci containing high-confidence fine-mapped Neanderthal variants, the observed enrichments provide complementary evidence that Neanderthal introgressed regions contribute to cancer risk beyond the genome-wide significant loci captured in the fine-mapping analysis.

## DISCUSSION

Interbreeding between Neanderthals and humans emigrating from Africa *∼*47,000 years ago has left a complex phenotypic legacy among modern humans (Dannemann and Kelso, 2017; Simonti et al., 2016; McArthur, Rinker, and Capra, 2021; Wei et al., 2023). Given the effects of Neanderthal ancestry on immune and metabolic processes linked to cancer risk and progression, we integrated maps of introgression and cancer GWAS data to evaluate the contribution of Neanderthal alleles to germline risk for multiple cancers. We discovered that risk loci for multiple cancers in both European and East Asian ancestry groups harbor Neanderthal alleles. Fine-mapping revealed 32 associations likely caused by Neanderthal variants.

Given the small contribution of common germline variants to risk for most cancers, the Neanderthal associations may be somewhat surprising. Our analyses reveal several insights that help explain the presence of associations between Neanderthal variants and cancer. First, while 14 of the significantly cancer-associated Neanderthal variants increase risk, 18 Neanderthal variants decrease risk. This differs from most other traits studied where Neanderthal variants mostly increase risk and many contribute to their maintenance.

Second, a substantial number of the associations we discover (9 of 32) are in regions with evidence of adaptive introgression, including *BNC2* and *PABPC4*. At *BNC2*, the Neanderthal allele is associated with lighter pigmentation as well as with increased skin cancer risk (Table S12). Pigmentation levels govern cumulative ultraviolet exposure. This suggests a mechanism of mediated pleiotropy, in which the cancer association is a downstream consequence of the adaptive pigmentation effect rather than an independent functional effect. Several of the other adaptive introgression loci with cancer associations include genes like *PABPC4* that encode multi-functional proteins involved in viral interactions and immune response, further suggesting pleiotropic mechanisms underlying the cancer associations. Because most cancer risk is realized after reproduction, an allele favored for an earlier-life effect can carry a cancer cost that purifying selection acts on only weakly.

Finally, with the exception of breast cancer, Neanderthal variants are depleted or neutral in their contributions to nearly all tested cancers. This is consistent with patterns seen for most non-cancer traits due to the selection against Neanderthal ancestry in many functional regions of the genome. Several features of breast cancer may contribute to the enrichment. Breast cancer risk depends strongly on lifetime reproductive hormone exposure and sensitivity. Neanderthal haplotypes have been associated with reproductive phenotypes, e.g., via a change in the progesterone receptor that increases its sensitivity and increases fertility (Zeberg, Kelso, and Pääbo, 2020). Thus, it is possible that the enrichment reflects antagonistic pleiotropy in which variants retained for early life effects on reproductive success could contribute to increased breast cancer risk. Similarly, breast tumors develop in an environment dominated by adipocytes (fat cells) in which lipid-induced inflammation often contributes to risk. Neanderthal alleles have adaptive effects on lipid metabolism (Khrameeva et al., 2014) and immune activity (Abi-Rached et al., 2011; Mendez, Watkins, and Hammer, 2012; Sams et al., 2016; Dannemann and Kelso, 2017; Zhou et al., 2021; Jagoda et al., 2022; Zeberg and Pääbo, 2020; Zeberg and Pääbo, 2021). Finally, the genetic architecture of breast cancer is highly polygenic and focused in non-coding regulatory regions, and the Neanderthal variants that persist in modern human genomes are also predominantly regulatory rather than coding (McCoy, Wakefield, and Akey, 2017).

While our results provide an important first step toward understanding how introgression from Neanderthals has contributed to germline cancer risk in modern humans, we acknowledge several limitations to our approach. First, while cancers vary in their genetic architecture and germline risk, our results are influenced by differences statistical power between the cancer GWAS considered. The number of fine-mapped Neanderthal associations identified per GWAS correlates with statistical power. We also note that several of the GWAS considered either used the same cohort (e.g., basal and squamous cell carcinoma) or included data from the UK Biobank. For example, the basal and squamous cell carcinoma summary statistics are the output of a multi-trait analysis of GWAS across keratinocyte cancer phenotypes rather than single-trait association tests, so their effect estimates borrow information across those phenotypes. Thus, the two associations we report at *BNC2* may reflect a single signal. However, we note that the breast cancer GWAS are independent. Second, while we consider multiple introgression maps, they are ultimately incomplete and are also underpowered to detect certain classes of variants, including reintroduced ancestral alleles as exemplified by uncalled variants near *PTHLH*. Therefore, the posteriors reported here are likely lower bounds on the Neanderthal contribution. Third, the methods for identifying Neanderthal variants and for fine mapping depend on an external LD reference. The 1000 Genomes panels used here comprise roughly 500 individuals per ancestry group, so LD involving variants too rare to be well represented cannot be estimated reliably and those variants cannot be fine-mapped. The panel also stands in for each study cohort’s own LD, and mismatch between the two is an inherent limitation of summary-statistic-based fine-mapping. The kriging diagnostic flagged 520 variants as potentially inconsistent with the reference panel; only 12 fell within a credible set, none were Neanderthal variants, and none occurred among the Neanderthal-majority associations (Table S17), so this is unlikely to affect the associations we report. Fourth, despite the increasing availability of genetic data from peoples of non-European ancestry (Fatumo et al., 2022), there are few publicly available cancer GWAS beyond those used in the present analysis. As additional data become available, disentangling ancestry-group specific cancer risk and differences in Neanderthal contributions will be possible. This will be particularly relevant given known differences in Neanderthal ancestry between continental groups (Vernot and Akey, 2014). Further, additional data from individuals from the Americas, southern Asia, and Oceania will enable analysis of Denisovan ancestry on cancer risk, as Denisovan ancestry is highest in these populations (Vernot et al., 2016; Sankararaman et al., 2016; Browning et al., 2018).

Mapping the contribution of Neanderthal variants to modern human cancer risk adds to our understanding of phenotypic differences and similarities between these groups. However, it also provides valuable context for interpreting the causes of cancer and the pleiotropic mechanisms sustaining cancer-associated variation. Neanderthal variation can be dated and its selective history reconstructed, making it a tractable system for asking how early-life selection shapes the genetic architecture of late-onset disease.

## METHODS

### Cancer GWAS

We collected metadata on publicly available cancer GWAS summary statistics, primarily using those available from GWAS Catalog (Cerezo et al., 2025). In total, we considered 227 GWAS among 17 cancers whose discovery sample descriptions were “East Asian” or “European” (Table S1). We did not consider GWAS whose summary statistics were derived from multiple superpopulations (i.e., East Asian and European), because downstream analyses required reference panel data from a genetically similar panel.

Ultimately, we selected a single GWAS for each cancer and ancestry group combination, prioritizing those with a large overall discovery sample size and balanced ratio of cases to controls. This resulted in 12 GWAS among individuals genetically similar to East Asians and 17 GWAS among individuals genetically similar to Europeans (Table 1, Table S2). Cases in each study are defined by primary site. We used each contributing study’s phenotype definition without modification; these are reported in Table S2. Two differ from the cancer label used here: the European esophageal cancer GWAS reports esophageal or stomach cancer, which overlaps the phenotype of the European gastric cancer GWAS, and the East Asian liver cancer GWAS reports hepatic bile duct cancer, whereas the European liver cancer GWAS reports cancer of the liver and intrahepatic bile duct. The basal cell carcinoma and squamous cell carcinoma summary statistics (Seviiri et al., 2022), reported in Table S2 with an MTAG suffix, are the output of a multi-trait analysis of GWAS (MTAG; Turley et al., 2018) across keratinocyte cancer phenotypes rather than single-trait association tests. MTAG borrows power across genetically correlated traits, so the reported effect sizes are not independent of the other traits included in that analysis, and the effective sample size of the output exceeds the value implied by each cancer’s case and control counts. We computed effective sample sizes from the reported case and control counts throughout, for consistency with the other studies.

Summary statistics were downloaded from GWAS Catalog (Cerezo et al., 2025), except for breast cancer among European-like individuals, which was downloaded from the Breast Cancer Association Consortium website. We mapped rsIDs to summary statistics when needed using single-nucleotide variants (SNVs) present in gnomAD v2.1.1 (Karczewski et al., 2020) whose FILTER column was “PASS”, used liftOver (Hinrichs et al., 2006) where necessary to convert from hg38 to hg19 genomic coordinates, and calculated *β* from odds-ratios (ORs) when not provided using the following calculation: *β* = ln(OR).

### Neanderthal Introgression Data

Many methods have been developed to identify introgressed DNA in a target population, yet the sets of inferred introgressed variants and regions can vary substantially, even when applied to the same population (Chen, Velazquez-Arcelay, and Capra, 2026). Therefore, we used four different Neanderthal introgression datasets that span algorithm architectures and feature variant-level calls: Browning et al., 2018, Sankararaman et al., 2016, Skov, 2024, and Vernot et al., 2016.

Three of the four datasets below use at least one reference genome from an archaic hominin to identify variants and groups of variants or haplotypes. Hereafter, we refer to the latter as “regions”. The high-coverage (*∼*52X) Neanderthal genome used in the methods below is Denisova 5, an *∼*120,000 year old specimen from a female who was recovered from the Denisova Cave in the Altai Mountains of southern Siberia (Prüfer et al., 2014). We refer to this specimen as the “Altai Neanderthal”. Introgression methods may also identify Neanderthal alleles in modern humans using a second archaic hominin genome from a sister lineage to Neanderthals—Denisovans. A high-coverage (*∼*31X) genome is available from Denisova 3, a 70,000 year old specimen that was also recovered from Denisova Cave (Meyer et al., 2012). We refer to this specimen as the “Denisovan”.

The Browning et al. 2018 dataset was generated using Sprime—a scoring method that identifies introgressed haplotypes using an archaic reference genome, the expected length of such haplotypes, and clusters of derived alleles that are absent in an unadmixed outgroup. Sprime further adjusts the score based on the local mutation and recombination rates, allows for low-frequency introgression in the unadmixed outgroup and avoids windowing. To be conservative, we required that the variants in this dataset match the Altai Neanderthal (NMATCH = match). We defined introgressed regions per the authors such that a Neanderthal region must comprise *≥* 30 variants that could be assessed in both the Denisovan and Altai Neanderthal genomes (i.e., not filtered due to quality concerns). In addition, there must be an overall match rate *≥* 0.6 for Neanderthal alleles and *≤* 0.4 for Denisovan alleles. When applied to the non-African 1000 Genomes Project (Auton et al., 2015) subpopulations and Papuans from the Simons Genome Diversity Project or SGDP (Mallick et al., 2016), Sprime identified 258,948 variants and 1,337 regions.

The Sankararaman et al. 2016 dataset was generated using a conditional random field (CRF) to infer Neanderthal ancestry at individual single-nucleotide polymorphisms (SNPs) along modern human haplotypes. The model uses a panel of sub-Saharan African Yoruba (YRI) haplotypes, which are assumed to harbor no Neanderthal ancestry, together with a high-coverage Neanderthal genome to characterize the observed sequence data and infer the unobserved Neanderthal ancestry state. The model accounts for the genetic distance between adjacent SNPs and uses the observed data to estimate the marginal probability of Neanderthal ancestry at each SNP. We used the first of two available datasets, where haplotypes are inferred using either the Denisovan or Altai Neanderthal as reference and an African genome as the unadmixed reference. We did not apply any quality filtering to the provided haplotypes or “regions” but required that variants had a mean posterior probability of Neanderthal origin > 0.5. When applied to non-African SGDP samples (Mallick et al., 2016), the CRF identified 892,930 variants and 3,791 regions.

The Skov 2024 dataset was generated by applying a Hidden Markov Model to modern human sequences. This model has two hidden states: 1) ingroup or segments of non-introgressed DNA and 2) archaic or segments of introgressed DNA. This method leverages the density of private variants in a queried sample, such that segments of introgressed DNA will have a longer time to accumulate variants due to deeper coalescence, and thus will exhibit a higher density of private variants compared to segments of non-introgressed DNA. This method does not require an archaic genome for comparison but does require an unadmixed outgroup (i.e., Africans). We identified regions by filtering for segments whose mean posterior probability that a segment was archaic was *≥* 0.8 and whose likely source was either Neanderthal or both (i.e., Neanderthal and Denisovan). We collated segments called in 1000 Genomes and Human Genome Diversity Project (Bergström et al., 2020) samples, merged intervals using bedtools (Quinlan and Hall, 2010), and used liftOver (Hinrichs et al., 2006) to change coordinates from the hg38 to hg19 human reference assembly. We identified variants by filtering for those that were shared with at least one of the following Neanderthal genomic samples: Altai, Chagyrskaya, Forbes, Goyet, HST, LesCottes, Mez1, Mez2, Scladina, Spy, Vindija, VindijaG1. When applied to samples from 1000 Genomes and HGDP, the HMM identified 354,656 variants and 4,459 regions.

The Vernot et al. 2016 dataset was generated using S*, which identifies candidate introgressed haplotypes based on clusters of derived variants that are absent from an unadmixed reference population and occur in high linkage disequilibrium. Introgressed haplotypes are then identified by maximizing the sum of scores among all SNP subsets at a particular locus. Tag SNPs are those variants that match an archaic allele and occur with at least two other tag SNPs in a 50 kb window. Haplotypes were defined as regions encompassing *≥* 5 tag SNPs in LD within a given human population (*r*^2^ *≥* 0.8). We collated tag SNPs called in 35 individuals from the Bismarck Archipelago as well as individuals from the East Asian, European, and South Asian 1000 Genomes superpopulations. We merged overlapping haplotypes using bedtools (Quinlan and Hall, 2010) to be conservative when calling regions. Heterozygous Neanderthal allele calls were set to missing. This procedure yielded 232,262 variants and 6,838 regions.

We merged the variant calls from the four datasets into a single union with one row per hg19 position, retaining any position called by at least one map (N = 1,085,232). Where maps reported different rsIDs or different allele pairs at a position, we retained the first value in map order (Browning, Sankararaman, Skov, Vernot) and recorded the disagreement. Where maps reported different Neanderthal alleles at a position, or where the reported Neanderthal allele was not one of the variant’s two alleles, the Neanderthal allele was set to missing.

To assess how common Neanderthal variants are in present-day populations, we retrieved allele frequencies for every variant in the union with an rsID from gnomAD v4.1.0 genomes (Chen et al., 2024), using PASS-filtered SNVs and matching variants by rsID, since gnomAD v4.1 is on GRCh38 and the introgression maps are on hg19. We report the minor allele frequency (MAF), the frequency of the rarer allele, in the African/African American, East Asian, non-Finnish European and South Asian gnomAD groups, and used MAF > 1% as the threshold at which variants are common enough to be detectable in GWAS of these sizes (Fig. S2).

### Identifying Cancer GWAS Loci with Neanderthal Variants

We constructed separate LD reference panels using 1000 Genomes data for East Asian (EAS) and European (EUR) samples. We excluded the HG01783 sample from the EUR reference panel because this individual exhibits genetic affinity for both Iberian populations in Spain and Mende populations from Sierra Leone. This resulted in 504 and 502 samples representing the EAS and EUR superpopulations, respectively. We used bcftools, version 1.21 (Li, 2011), to subset biallelic SNVs per superpopulation using “view -m2 -M2 -v snps” and plink2, version 2.0.0-a.6.9 (Chang et al., 2015), to generate the files required for LD calculation. We retained the first occurrence of duplicate variants.

Next, we retrieved significant variants from the summary statistics of each cancer GWAS using the standard genome-wide significance threshold (*P* = 5 *×* 10^−8^). Introgressed variants were identified by matching each significant GWAS variant against the union of the four introgression maps, first by rsID and then, for variants that remained unmatched, by hg19 position. A position match was retained only when the two alleles reported by the introgression maps agreed with the GWAS effect and non-effect alleles, or when no map reported alleles at that position. We performed LD clumping using plink, version 1.90b6.26 (Purcell et al., 2007), and ran the procedure at three *r*^2^ thresholds: 0.2, 0.5, and 0.8. We set “clump-p1” and “clump-p2” to 1 because only genome-wide significant variants were included in the clumping procedure and used the default “clump-kb” value of 250. We conducted this procedure separately on the introgressed subset and on all significant GWAS variants. For each GWAS, we defined loci as non-singleton clumps, such that at least two significant variants occurred in LD at the specified *r*^2^ threshold. We calculated the proportion of GWAS loci containing introgressed variants by dividing the number of loci identified from the introgressed-variant subset by the total number of loci identified from all significant variants.

### Fine-Mapping

To conduct fine-mapping, we first identified loci in each GWAS centered on genome-wide significant Neanderthal variants. We used a greedy approach by p-value, identifying the most strongly associated variant as the index variant for the first locus. All Neanderthal variants within 500 kb were assigned to that locus. Then, the next most strongly associated remaining variant defined the next. We fine-mapped a *±*500 kb window around each index variant with SuSiE-RSS from the susieR package (Zou et al., 2022). For each locus, we used the aforementioned ancestry-group-specific LD reference panels calculated from 1000 Genomes. We removed variants with missing rsIDs, duplicated rsIDs, or standard errors that were missing or *≤* 0. Variants monomorphic in the relevant reference panel were removed with PLINK’s “–mac 1”, which retains singletons and is equivalent to a minor allele frequency threshold of 1/(2*N*), where *N* is the number of individuals in that ancestry group in 1000 Genomes. Strand-ambiguous variants (A/T and C/G) were excluded before alignment. Variants were aligned to the corresponding LD reference panel by effect and non-effect alleles, including reverse-complement strand matching, and effect estimates were sign-flipped when necessary. Variants with irreconcilable allele mismatches were excluded. GWAS effect estimates and standard errors were used to calculate Z-scores as *Z* = *β*/*SE*. PLINK was run with “–keep-allele-order” both when extracting the panel and when computing correlations, so that the signs of the LD estimates correspond to the A1 alleles recorded in the .bim file used to align effect estimates. The LD matrix was calculated as pairwise Pearson correlations among variants in the ancestry-specific reference panel. Missing correlations were set to zero, the matrix was symmetrised, 10^−4^ was added to the diagonal for numerical regularization and to ensure the matrix was positive definite, and the result was converted back to a correlation matrix.

We assessed consistency between the GWAS summary statistics and the ancestry-specific LD reference in the initial fits using the kriging RSS diagnostic implemented in susieR. Variants with log *LR* > 2 and |*z*| > 2 were flagged as potentially inconsistent with the LD reference (Table S17). We identified 520 variants across 115 loci, of which one exceeded the severe threshold of log *LR* > 10; that variant belongs to no credible set. Twelve of the 520 fell within a credible set; none is a Neanderthal variant, none belongs to a genome-wide significant credible set, and none falls within any of the 32 Neanderthal-majority associations.

We set the GWAS sample size as the effective sample size, defined as

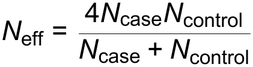

We used a maximum of 1,000 iterations, an *L* = 10, a minimum *λ* of 0.01, and a convergence tolerance of 10^−3^. The residual variance was estimated using the susie_rss default. We allowed the prior variance of each single-effect component to be estimated from the data. We initially did not perform model refinement. Credible sets were constructed using the default 95% coverage and a purity (minimum absolute correlation among variants within each credible set) of 0.5. For each locus we retained the original fit if it converged; otherwise a refined refit that converged, preferring *L* = 5 over *L* = 10. Loci with no converged fit were considered unresolved. A locus whose refit disagreed with the original about which genome-wide significant variants carried the signal (Jaccard < 0.3; one locus, at 0.25) was also treated as unresolved, since convergence to an answer that depends on initialization is not a resolved locus.

After fine-mapping, we filtered for credible sets that contained at least one variant reaching genome-wide significance (|Z| *≥* 5.45, *P* = 5 *×* 10^−8^). We merged credible sets from different index windows within the same GWAS when they shared at least half their variants (Jaccard *≥* 0.5). Next, we annotated whether or not credible set variants were Neanderthal by presence in the merged union of the four introgression maps. Finally, we calculated the posterior probability that the causal variant is Neanderthal by summing the posterior inclusion probability of every Neanderthal variant in the fine-mapped window, not only those within the credible set. We designated Neanderthal-majority associations as those where Neanderthal posterior probability was > 50%.

We assigned a direction of effect to each Neanderthal-majority association. For each Neanderthal variant we oriented the GWAS effect size to the Neanderthal allele, taking *β* as reported when the Neanderthal allele was the effect allele and –*β* when it was the non-effect allele. Three of the four introgression maps report a Neanderthal allele; the Sankararaman et al. 2016 conditional random field infers Neanderthal ancestry at a SNV without naming an allele. For variants called only by that map, we took the Neanderthal allele to be the derived allele, determined from the Ensembl human ancestor sequence (GRCh37, EPO alignment; Herrero et al., 2016). Reintroduced ancestral alleles were excluded from this assumption, since at those sites the Neanderthal allele is ancestral by definition. Each association was then assigned the direction carrying the greater summed posterior inclusion probability across its Neanderthal variants.

We performed conditional analysis at the *PTHLH* locus. We conditioned both the East Asian and European breast cancer windows at this locus on the strongest association signal in the European GWAS: rs7297051. Z-scores were computed as 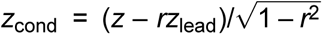. We defined the attenuation ratio as the conditional |*Z*| divided by the marginal |*Z*| for the credible set’s lead variant (highest PIP). To assess power, we rescaled the East Asian *Z* scores by the ratio of *Z* per unit effect size at rs788464 (a variant in both credible sets) between the two GWAS, and recomputed the conditional statistic holding *r*, the correlation with rs7297051 in the matched reference panel, fixed.

### Structural Variants

To assess whether structural variants (SVs) might mediate the Neanderthal-associated signals at *BNC2* and *PTHLH*, we queried the SVatalog database (Chirmade et al., 2026), which provides LD estimates between SNPs and structural variants from 1000 Genomes long-read sequencing data. For each locus, we extracted all SVs within the fine-mapped region and computed *r*^2^ and *D* with the lead Neanderthal credible set variant. We additionally queried gnomAD-SV v4 (Collins et al., 2020) for common structural variants (AF > 0.01) within 250 kb of the Neanderthal credible set variants for both loci. At the *PTHLH* locus, we further examined 578 phased haplotypes from long-read genome assemblies spanning chr12:27,700,000–28,100,000 (hg19). Haplotype sequences were classified by total interval size relative to the T2T reference; insertion breakpoints were localized using k-mer anchoring, and hallmarks of retrotransposition (poly-A tail, target site duplication) were assessed from the extracted insertion sequence.

### Functional Annotations

We leveraged multiple datasets to assess functional effects of fine-mapped variants. To label each Neanderthal-majority association with a gene, we used protein-coding and lncRNA genes from GENCODE v50 (basic set, GRCh37 mapping) and assigned the gene whose body is closest to the credible set’s highest-PIP variant, with a distance of zero when the variant falls within the gene. We retrieved adaptively introgressed regions from Iasi et al., 2026 (their Table S26). We used those regions labelled as “Regions of high frequency in both ancient and present-day (PD) samples” and “Regions of low frequency in ancient and high in present day (PD) samples”. For each Neanderthal-majority association, we defined the credible set span as the interval from its first to its last variant and used bedtools closest (Quinlan and Hall, 2010), through pybedtools, to find the nearest adaptively introgressed region and the distance to it; a distance of zero indicates overlap (Table S9). Gene models shown in the locuszoom panels are from the same annotation; they are limited to two rows, so some non-coding models in dense windows are not shown. We retrieved candidate cis-regulatory elements (cCREs) as profiled by ENCODE 3, using the global set across all cell lines and tissues (Abascal et al., 2020), and lifted them to hg19. We retrieved reintroduced ancestral allele calls from Rinker et al., 2020.

### eQTL Analysis

We first queried the GTEx Portal (GTEx v10) (The GTEx Consortium, 2020) for expression quantitative trait loci (eQTLs) and splicing QTLs (sQTLs) among the credible set variants at each of the three example loci (*BNC2*, *SLC24A4* and *PTHLH*). None of the *BNC2* credible set variants is a significant eQTL or sQTL in any tissue. For loci with significant associations, we retrieved eQTL results through the GTEx Portal API for rs788464 and rs10843134 at *PTHLH* and rs79342051 at *SLC24A4*. For each variant, we retrieved every gene–tissue pair in which it is a significant single-tissue eQTL, as defined by GTEx, together with the normalized effect size (NES), its *P*-value and per-sample normalized expression by genotype. For selected genes (*PTHLH*, *CCDC91*, ENSG00000247934 and *SLC24A4*), we also retrieved each variant’s effect in every tissue, whether or not it was significant. GTEx reports effects for the alternate allele; we oriented all effects and genotypes to the allele of interest at each variant (the Neanderthal allele, or the reintroduced ancestral allele at rs788464). Standard errors were computed as NES divided by the reported *t* statistic. Colocalization was not tested.

### Associations with Other Traits

To identify other phenotypes associated with the Neanderthal haplotypes at *BNC2* and *SLC24A4*, we queried Open Targets Platform (Buniello et al., 2025) for every variant in our cancer credible sets at these loci (Tables S8 and S12), after converting positions from hg19 to GRCh38 using liftOver (Hinrichs et al., 2006). We retained all Open Targets credible sets containing at least one of these variants and recorded the posterior probability of the query variant, along with each credible set’s study, reported and mapped trait, lead variant, lead-variant effect size and *P*-value, fine-mapping method and top locus-to-gene (L2G) prediction.

Because Open Targets reports effect sizes for the alternate allele of each credible set’s lead variant, we oriented these effects to the Neanderthal haplotype. Lead variants present in our cancer credible sets were oriented by comparing the sign of their cancer effect with that of the Neanderthal allele. For the remaining lead variants, we determined which allele is inherited with the Neanderthal allele of rs2026805 (*BNC2*) using the LDproxy tool from LDlink (Machiela and Chanock, 2015) in the 1000 Genomes Project Phase 3 European (EUR) panel. Lead variants in weak linkage disequilibrium with the Neanderthal haplotype (*r*^2^ < 0.8) or located more than 1 Mb from it were treated as independent signals and excluded from interpretation.

### Neanderthal Allele Frequency Through Time

rs2026805, the lead variant of the *BNC2* credible sets, is not reliably genotyped across ancient samples. We therefore used AncientProxy (Brand and Capra, 2026) to identify rs2026804 as a proxy (*r*^2^ = 0.972 in TOPMed Europeans) and obtained its genotype calls for the Allen Ancient DNA Resource (AADR) v54.1.p1 from the same resource. The allele counted is the C allele at rs2026804, which is the Neanderthal allele reported by Skov, 2024 and the derived allele relative to the human–chimpanzee ancestor. Archaic hominin genotypes were excluded, so all frequencies reported are among modern humans.

Where a specimen was represented by more than one library, a single genotype was retained, preferring diploid shotgun calls. These are the only source in this dataset able to call a heterozygote: every heterozygous call comes from a diploid shotgun library and no other source produces one. The remaining sources are pseudo-haploid, reporting a true heterozygote as one homozygote or the other, and each therefore contributes a single allele rather than two. Among pseudo-haploid sources the preference order is arbitrary and only breaks ties where two libraries both carry a call.

Specimens were binned by mean date into [50-20), [20-10), [10-5), [5-2) and [2-0) ka, with wider bins at greater ages because sample density falls off; bins include the older bound, so a specimen dated exactly 2 ka falls in [2-0). Present-day samples were held out as a separate category rather than folded into the youngest bin, because they are a worldwide anthropological panel rather than a sample of any one population and their regional frequencies are therefore not comparable to the ancient bins or to continental frequencies from gnomAD (Karczewski et al., 2020) or 1000 Genomes. Within each bin and AADR region we report the derived allele frequency and a 95% Wilson score interval, which is used in place of the normal approximation because the latter has zero width where no derived allele is observed, as in the oldest bin.

### Heritability Enrichment

Heritability enrichment was estimated with LDSC (Bulik-Sullivan et al., 2015). The East Asian colorectal cancer GWAS was excluded because its summary statistics report no effect sizes, leaving 28 studies. Summary statistics were formatted against the ancestry-matched HapMap3 variant list. For each introgression map and ancestry group we generated annotation files and LD scores from the ancestry-matched 1000 Genomes PLINK filesets. Each map was then modelled separately as a single annotation added to the ancestry-matched baselineLD v2.2 model (Gazal et al., 2017), with ancestry-matched regression weights and 1000 Genomes allele frequencies, giving 28 *×* 4 estimates. Heritability is reported on the observed scale, since population prevalences were not available for these studies; observed-scale estimates depend on each study’s case fraction and are not comparable between cancers. The basal and squamous cell carcinoma summary statistics were genomic-control corrected by their authors (*λ* = 1.286 and 1.172), which deflates their reported intercepts and heritabilities, and are MTAG rather than single-trait output; enrichment is a ratio of proportions and is less sensitive to both, though MTAG test statistics are not those of a single-trait GWAS. Several studies include UK Biobank participants and are therefore not fully independent, though neither breast cancer GWAS does. Enrichments were interpreted only for GWAS with enough genome-wide signal to support them: mean *χ*^2^ *≥* 1.02 and an observed-scale heritability *≥* 0.05 (Tables S11-12). Enrichment *P*-values were corrected for multiple testing across the 32 tests conducted among the GWAS passing heritability QC (eight GWAS *×* four introgression maps) using the Benjamini–Hochberg procedure.

We also estimated heritability enrichment for breast cancer from individual-level genotypes in the UK Biobank (application 226956) using GENIE v1.2.1 under an additive genetic model without an environment interaction term (Pazokitoroudi et al., 2024). Cases were the 14,346 individuals with a breast cancer diagnosis (ICD-10 code C50) in data field 40006. Controls were individuals with no cancer diagnosis recorded in that field; we selected three per case with nearest-neighbor matching in MatchIt (Ho et al., 2011) on sex, age, Townsend Deprivation Index and the top 20 genetic ancestry principal components, and these 23 variables were also included as fixed-effect covariates. Genotypes were filtered to variants with MAF > 0.01, missingness < 0.01 and Hardy–Weinberg equilibrium *P ≥* 10^−7^, leaving 532,664 variants across the autosomes and chromosome X for 54,472 individuals. In contrast to the LDSC analysis, the four Neanderthal introgression maps were modelled jointly, as four annotations (4,933, 14,120, 5,844 and 2,965 variants for the Browning, Sankararaman, Skov and Vernot maps, respectively) alongside a fifth annotation comprising the remaining 516,376 variants. Because the maps overlap, we report GENIE’s overlapping-annotation enrichment estimates. Standard errors were estimated from 100 jackknife blocks. Total observed-scale heritability was 0.034 (SE 0.005).

### Analysis

All data analyses were performed using Bash, Python, and R scripts, some of which were implemented in Jupyter notebooks. All reported p-values are two-tailed, unless noted otherwise.

### Visualization

Results were visualized using Inkscape, version 1.1 (Inkscape Project, 2020) and ggplot2, version 4.0.3 (Wickham, 2016) implemented in R, version 4.5.3 (R Core Team, 2020).

## Data and Code Availability

We used publicly available data for most analyses. Two exceptions include analysis of long-read sequences near *PTHLH*, which were shared by Charikleia Karageorgiou and Omer Gokcumen (personal communication), and the heritability enrichment replication, which was performed using the UK Biobank (application 226956). Cancer GWAS were retrieved from GWAS Catalog or the relevant data hosting repository. Table S2 features URLs for each cancer GWAS summary statistics file. Browning et al. 2018 introgressed variant data were retrieved from https://data.mendeley.com/datasets/y7hyt83vxr/1. Sankararaman et al. 2016 introgressed variant data were retrieved from https://drive.google.com/drive/folders/1DyhMw0E1mXQUDNeGQbQrcQbHe0uTyK2h. Skov 2024 introgressed variant data were retrieved from https://zenodo.org/records/14136628. Vernot et al. 2016 introgressed variant data were retrieved from https://drive.google.com/drive/folders/0B9Pc7_zItMCVM05rUmhDc0hkWmc?resourcekey=0-zwKyJGRuooD9bWPRZ0vBzQ. 1000 Genomes Project Phase 3 data were retrieved from The International Genome Sample Resource (IGSR) (https://ftp.1000genomes.ebi.ac.uk/vol1/ftp/). gnomAD v2.1.1 and v4.1.0 genomes data were retrieved from https://gnomad.broadinstitute.org/data. gnomAD v4.1 structural variant data were queried from the gnomAD browser (https://gnomad.broadinstitute.org); structural variant–SNP linkage disequilibrium data were obtained from SVatalog (https://svatalog.github.io/), both on November 6, 2026. The Iasi et al. 2025 adaptive introgression data were retrieved from https://www.science.org/doi/suppl/10.1126/science.adq3010/suppl_file/science.adq3010_supplementary_tables.zip, Table S26. Gene models were retrieved from GENCODE release 50, GRCh37 mapping, Basic set (https://ftp.ebi.ac.uk/pub/databases/gencode/Gencode_human/release_50/GRCh37_mapping/gencode.v50lift37.basic.annotation.gff3.gz). ENCODE 3 cCREs were retrieved from https://downloads.wenglab.org/V3/GRCh38-cCREs.bed. eQTL and sQTL data are from GTEx v10 and were accessed through the GTEx Portal (https://gtexportal.org/home/) and its API (https://gtexportal.org/api/v2/) on November 1, 2026. Credible sets for other traits were retrieved from the Open Targets Platform, release 26.09 (https://platform.opentargets.org/) on November 1, 2026. LDlink was accessed from https://ldlink.nih.gov/ on November 1, 2026. Ancient DNA genotype calls for rs2026804 were obtained from the AncientProxy catalog (Brand and Capra, 2026, https://github.com/brandcm/AncientProxy), which derives them from the Allen Ancient DNA Resource v54.1.p1 (Mallick et al., 2024; Harvard Dataverse, https://doi.org/10.7910/DVN/FFIDCW). Reintroduced ancestral alleles were retrieved from https://github.com/DaRinker/Neanderthal.reintroduction/blob/master/Rinker_Suppl.File1.tar.gz. Stratified LD score regression data were retrieved from https://zenodo.org/records/10515792. All code used to conduct analyses and generate figures is publicly available on GitHub (https://github.com/brandcm/neanderthal_ancestry_and_cancer_risk) and will be archived upon publication. Larger data files not housed in the GitHub repository can be accessed here:https://zenodo.org/records/24015068.

## Acknowledgements

We thank Charikleia Karageorgiou and Omer Gokcumen, who generously shared data from long-read assemblies. We are grateful to Jon Akutagawa, Iona Cheng, Haruna Tomono, Mimi Ton, and Elad Ziv, who offered thoughtful suggestions throughout this project. We also thank members of the Capra Lab who gave helpful feedback. We are grateful for the support and resources from the Wynton High Performance Compute Cluster at University of California San Francisco. CMB was funded by National Institutes of Health grant 5T32CA112355-19. JAC and CMB were funded by National Institutes of Health grant R35GM127087.

## Author Contributions

Conceptualization, CMB and JAC; Formal Analysis, CMB and YC; Writing – Original Draft, CMB, YC, and JAC; Writing – Review & Editing, CMB, YC, and JAC.

