## Supplementary Information for "The contribution of Neanderthal ancestry to cancer risk"

#### Supplementary Text

##### The *PTHLH* Locus

Fine-mapping identified Neanderthal-majority associations for breast cancer at *PTHLH* (chromosome 12) in both the East Asian and European GWAS (Fig. 5, Table S6). The European GWAS yields three credible sets at this locus: credible set 1 (CS1), a single variant (rs7297051); CS2, nine variants led by rs10843134; and CS3, 28 variants led by rs788464. The East Asian GWAS yields a single credible set of 34 variants led by rs1267211, which shares rs788464 with European CS3. Below we assess structural variants at this locus, the independence of these signals, the reintroduced ancestral alleles (RAAs) they contain, and their regulatory annotation.

##### Structural Variants

No common structural variant (SV) in the SVatalog database was in meaningful linkage disequilibrium with the credible set variants at *PTHLH* (maximum $r^2$ = 0.23 between P0000139595 and the non-Neanderthal credible set variant rs7297051; Table S14). The low $r^2$ despite perfect $D'$ reflects a large allele frequency mismatch (SV AF = 0.10 vs. SNP AF = 0.34), indicating the SV occurs on a subset of rs7297051-carrying haplotypes. No common, frequency-compatible structural variant was identified in gnomAD-SV v4 within 250 kb of the Neanderthal credible set variants (Table S13). However, examination of 578 phased haplotypes from long-read genome assemblies revealed a polymorphic full-length LINE-1 element (mean size 5,894 bp; range 5,620–6,541 bp) inserted in antisense orientation at chr12:27,945,230 (hg19), with a 15-bp target site duplication. The LINE-1 insertion allele frequency was 52.2%, substantially higher than the Neanderthal haplotype frequency (10% in Europeans), and the element is located approximately 195 kb upstream of the nearest Neanderthal credible set variant. Two gnomAD-SV v4 calls at this position — a 2.96-kb deletion (DEL_CHR12_35F6D990, AF = 0.047) and a complex SV (CPX_CHR12_C76181E1, AF = 0.029) — likely represent short-read artifacts of this LINE polymorphism. Given the allele frequency mismatch and physical distance from the credible set, this LINE-1 element is unlikely to mediate the Neanderthal-associated cancer risk signal at *PTHLH*.

##### Conditional Analysis

Because the European credible sets lie within 310 kb of one another, they may reflect a single underlying signal spread across variants in LD. Therefore, we assessed the independence of the three European associations by conditioning variants in both GWAS on the most strongly associated variant in the European GWAS: rs7297051 (Fig. S9). The maximum conditional $|Z|$ in CS2 and CS3 is 8.51 and 6.60, respectively, above the genome-wide significance threshold ($|Z| \geq 5.45$, $P = 5 \times 10^{-8}$). This indicates that there are three independent associations in the European GWAS. The same rs7297051 variant occurs at 22.3% frequency in Phase 3 1000 Genomes Project samples from the East Asian continental group (Auton et al., 2015). Conditional Z-scores in the East Asian GWAS do not meet genome-wide significance ($|Z|$ = 2.7–3.5). The two associations are also more tightly correlated in East Asians: rs7297051 and the East

Asian credible set variants have $|r| \approx 0.85$ in the East Asian reference panel, compared with $\approx 0.60$ between rs7297051 and the CS3 variants in the European panel, leaving less information with which to separate them. Rescaling the East Asian statistics to European power using the ratio of $Z$ per unit effect size (1.74x) at rs788464, a variant shared by both credible sets, yields a conditional $|Z|$ of 6.1 for the East Asian credible set, above the genome-wide threshold. This rescaling assumes equal effect sizes across the two GWAS and holds the LD estimates fixed; it indicates the magnitude of the power difference rather than constituting a formal test. However, the attenuation ratio—the fraction of marginal association that survives conditioning—is similar for the East Asian credible set and CS3 (0.44 and 0.43). This comparable ratio suggests that rs7297051 tags a second, unresolved association for breast cancer risk in the East Asian GWAS.

#### Reintroduced Ancestral Alleles

Six of the seven RAAs in the European credible sets also occur in East Asians; rs788464 is not identified as an RAA in East Asians. This may be due to introgression detection methods being least able to detect reintroduced ancestral alleles. Thus, the Neanderthal posterior probabilities reported at this locus are likely lower bounds.

#### Regulatory Annotation

Seven distinct credible set variants at this locus intersect cCREs, including rs7297051 from European CS1 and rs805513, a reintroduced ancestral allele shared by CS3 and the East Asian credible set; both fall within distal enhancer-like elements. The strongest regulatory evidence comes from CS2, whose lead variant, rs10843134, is an eQTL for *CCDC91* and the lncRNA ENSG00000247934 in breast tissue, with the Neanderthal allele increasing *CCDC91* expression; it is also an eQTL for *PTHLH* in esophagus (mucosa) (Figs. S10, S11). The reintroduced ancestral allele at rs788464 in CS3 increases expression of *PTHLH* in skeletal muscle (Fig. S10) and of ENSG00000257042 in adipose tissue, and decreases expression of ENSG00000247934 in cultured fibroblasts (Fig. S10). The two independent signals therefore converge on ENSG00000247934.

### Supplemental Figures

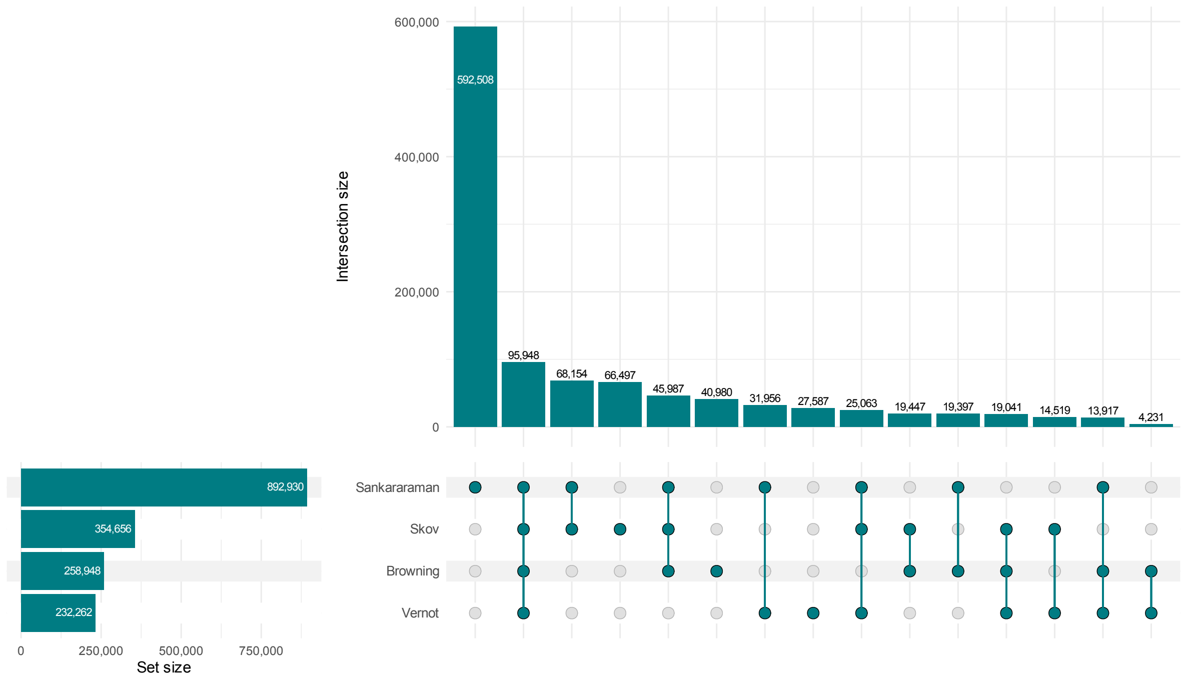


**Fig. S1. Overlap of Neanderthal introgressed variant calls across four published maps.** Horizontal bars give the total number of autosomal variants called by each map (Browning et al., 2018; Sankararaman et al., 2016; Skov, 2024; Vernot et al., 2016). The dot matrix enumerates combinations of maps, and vertical bars give the number of variants called by exactly that combination and no other; the vertical bars therefore partition the union of all calls (N = 1,085,232) rather than overlapping. Variants were matched across maps by hg19 position. Maps are ordered by the number of variants they call, and vertical bars by decreasing intersection size.

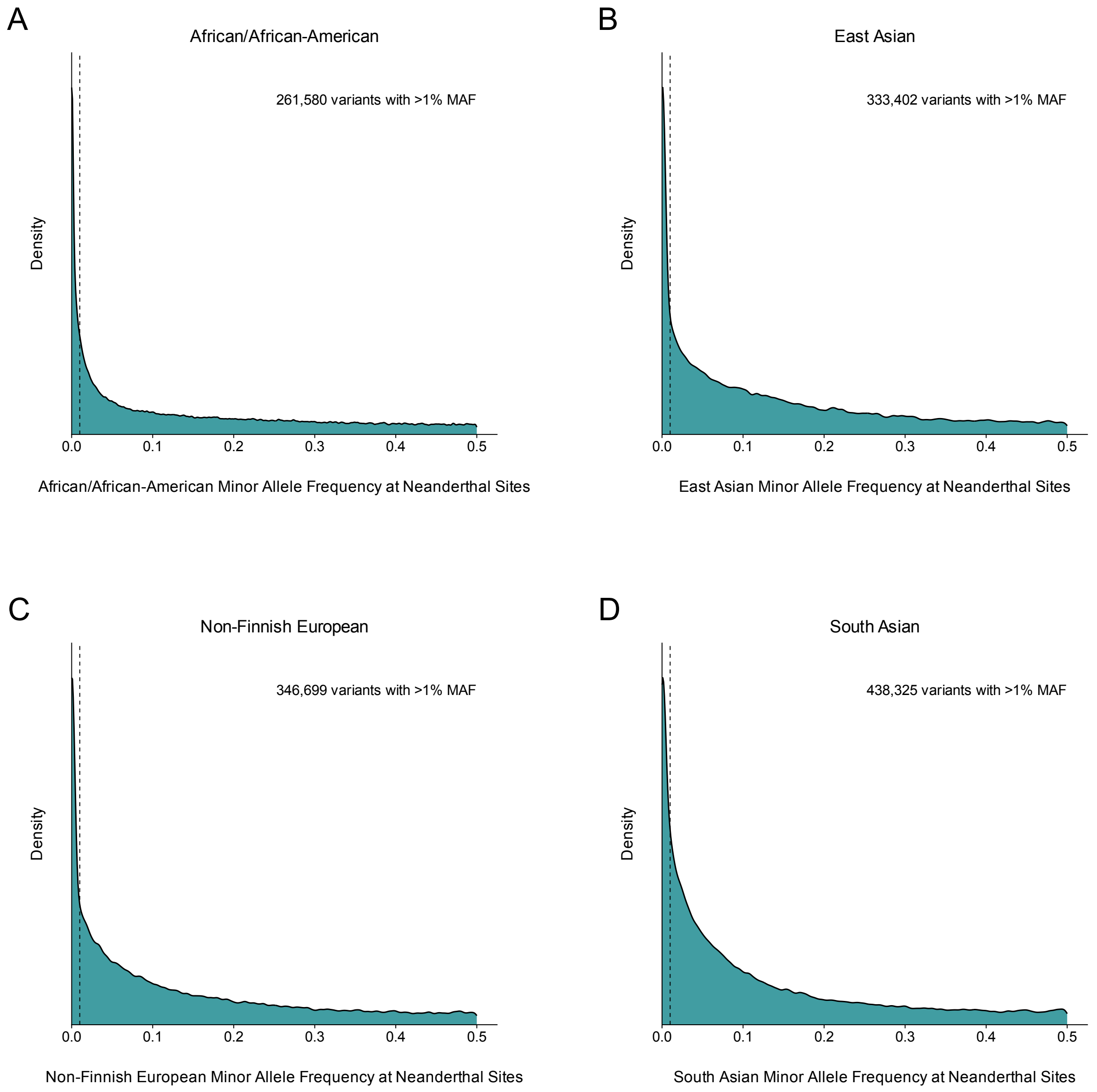


**Fig. S2. Neanderthal variants segregate at appreciable frequencies across different gnomAD ancestry groups.** Minor allele frequency distributions at sites harboring a map-called Neanderthal variant across gnomAD v4.1.0 ancestry groups: African/African-American (A), East Asian (B), Non-Finnish European (C), and South Asian (D). Allele frequencies were retrieved for 912,978 high-quality (FILTER = PASS) single-nucleotide variants (SNVs) identified across one or more of four Neanderthal maps (Browning et al., 2018; Sankararaman et al., 2016; Skov, 2024; Vernot et al., 2016). Interbreeding between Neanderthals and modern humans postdates the most recent major dispersal out of Africa, so frequencies in the African/African American ancestry group are expected to be the lowest; the residual frequencies observed there reflect recent European admixture within that group, back migration into Africa, and map calls that are not of introgressed origin. Minor allele frequency is the frequency of the rarest allele at each site; at multi-allelic sites the reference frequency was computed as $1 - \sum \text{ALT AF}$ and compared against all alternate alleles. Density is shown on a square-root scale. The dashed line marks a minor allele frequency of 0.01, the threshold below which variants are typically excluded from GWAS.

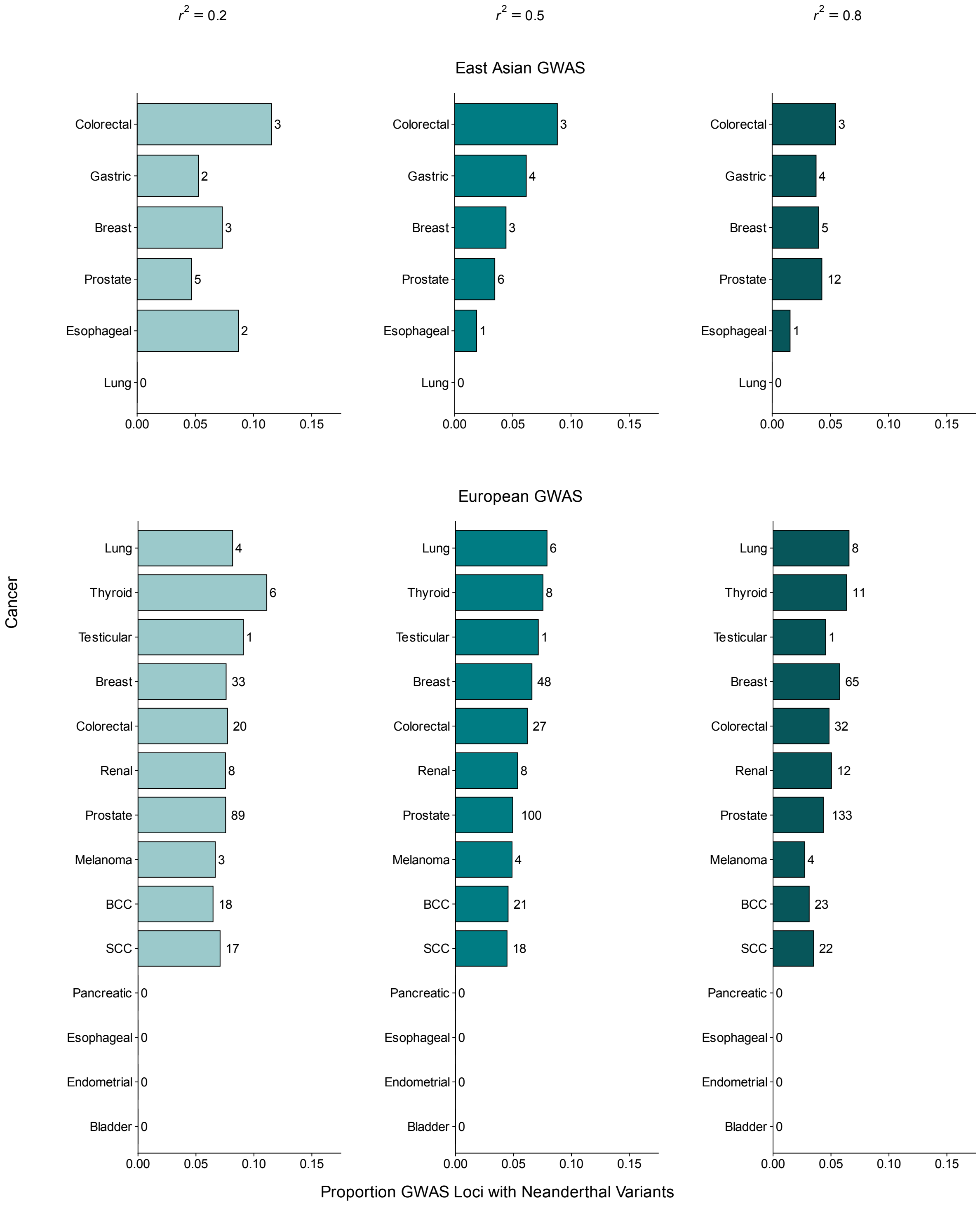


**Fig. S3. The proportion of GWAS loci with Neanderthal variants varies modestly across $r^2$ thresholds.** The proportion of independent cancer-risk loci harboring Neanderthal variants, stratified by ancestry group and $r^2$ threshold. Significant GWAS variants ($P < 5 \times 10^{-8}$) were LD-clumped within 250-kb windows using matching ancestry panels (East Asian or European) from 1000 Genomes. Loci were required to comprise at least two significant variants at the given $r^2$ threshold, since a locus resting on a single variant offers no secondary signals to assess; the same requirement was applied to Neanderthal variants. Within each ancestry group, cancers are ordered by the proportion at $r^2$ = 0.5. Bar labels give the number of loci harboring $\geq$ 2 Neanderthal variants, i.e., the numerator of each proportion. Bar shading denotes the $r^2$ threshold used for clumping.

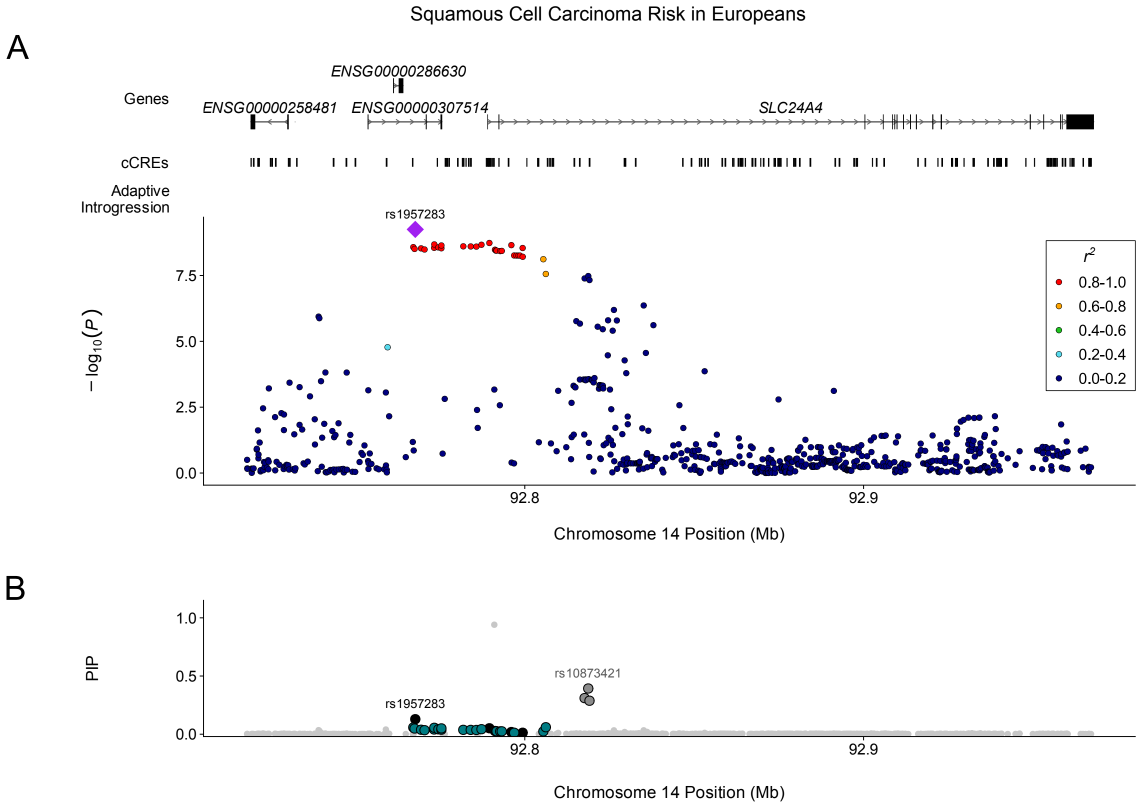


**Fig. S4. Neanderthal variants near *SLC24A4* associate with decreased risk of squamous cell carcinoma.**
(A) LocusZoom plot of variants from the Seviiri et al. (2022) squamous cell carcinoma GWAS conducted among individuals with genetic similarity to Europeans in 1000 Genomes. All variants in the summary statistics within the window are shown. Binned LD ($r^2$) with rs1957283 is indicated by color; variants absent from the 1000 Genomes reference panel, for which $r^2$ could not be calculated, are plotted in the lowest bin. The lead variant, rs1957283, is indicated by a purple diamond. cCREs, as profiled by ENCODE, are shown above. (B) Posterior inclusion probabilities (PIPs) for the variants in A. Credible sets are labelled CS1–CS*n* in the order reported by SuSiE at each locus. Teal and black mark members of CS1, the credible set this figure concerns, split by whether the variant is Neanderthal (teal) or not (black). No single variant exceeds PIP 0.13; the posterior is distributed across 21 Neanderthal variants in near-complete LD. Dark grey marks members of one other credible set at the locus (CS2) in the shown window, and light grey variants belong to no credible set.

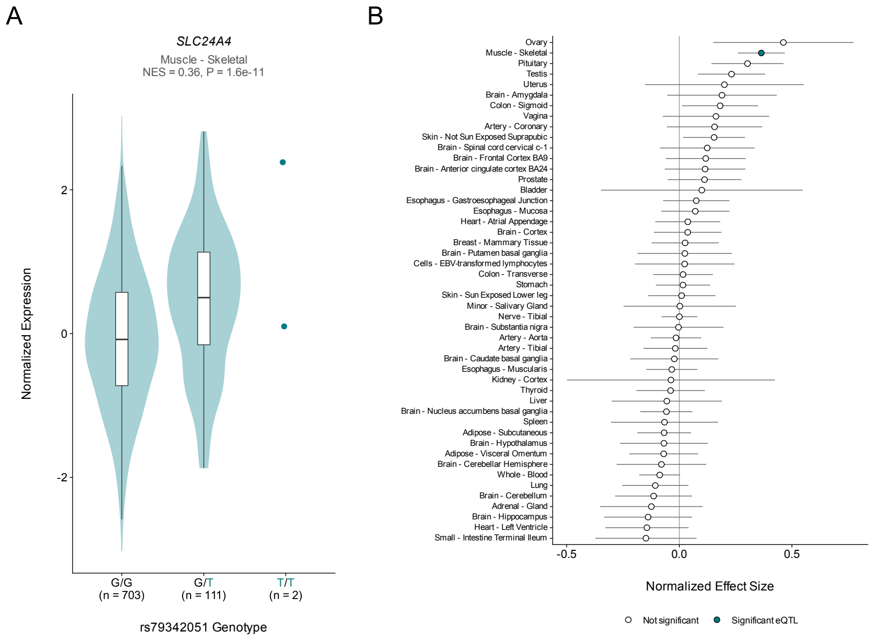


**Fig. S5. The Neanderthal allele at rs79342051 associates with increased *SLC24A4* expression in skeletal muscle.**
(A) Normalized expression of *SLC24A4* in skeletal muscle by rs79342051 genotype in GTEx. Violins show the distribution of per-sample expression for each genotype and boxes the median and interquartile range; genotypes with fewer than 10 samples are shown as individual points. Sample sizes are given below each genotype. The Neanderthal allele (T) is shown in teal. NES, normalized effect size of the Neanderthal allele (T). (B) NES of the Neanderthal allele (T) at rs79342051 on *SLC24A4* expression across GTEx tissues, with 95% confidence intervals. Filled points are significant eQTLs, as defined by GTEx; open points are not significant. Tissues in which GTEx did not test the variant are omitted.

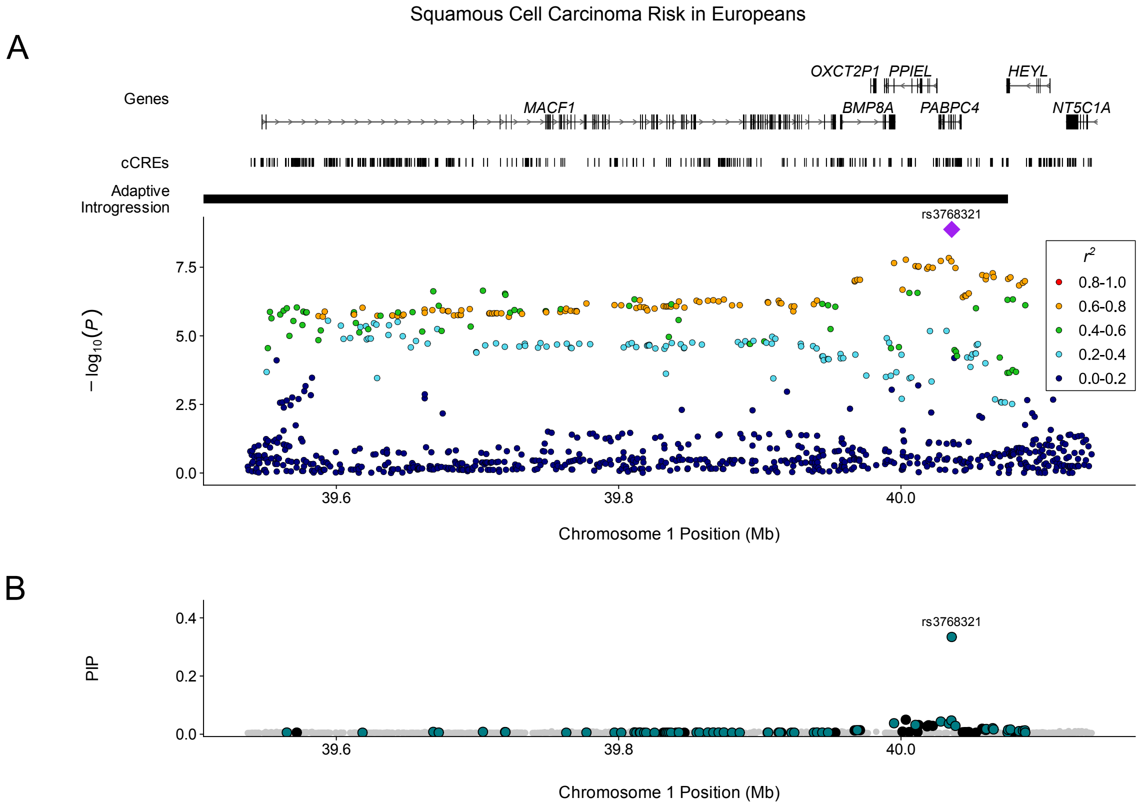


**Fig. S6. Adaptively introgressed Neanderthal variants on chromosome 1 associate with increased risk of squamous cell carcinoma.**

(A) LocusZoom plot of variants from the Seviiri et al. (2022) squamous cell carcinoma GWAS conducted among individuals with genetic similarity to Europeans in 1000 Genomes. All variants in the summary statistics within the window are shown. Binned LD ($r^2$) with rs3768321 is indicated by color; variants absent from the 1000 Genomes reference panel, for which $r^2$ could not be calculated, are plotted in the lowest bin. The lead variant, rs3768321, is indicated by a purple diamond. cCREs, as profiled by ENCODE, and adaptively introgressed regions from Iasi et al. (2025) are shown above. (B) Posterior inclusion probabilities (PIPs) for the variants in A. The credible set comprises 85 variants with purity 0.58; no single variant exceeds PIP 0.33. Teal and black mark members of CS1, the credible set this figure concerns, split by whether the variant is Neanderthal (teal) or not (black). Light grey variants belong to no credible set.

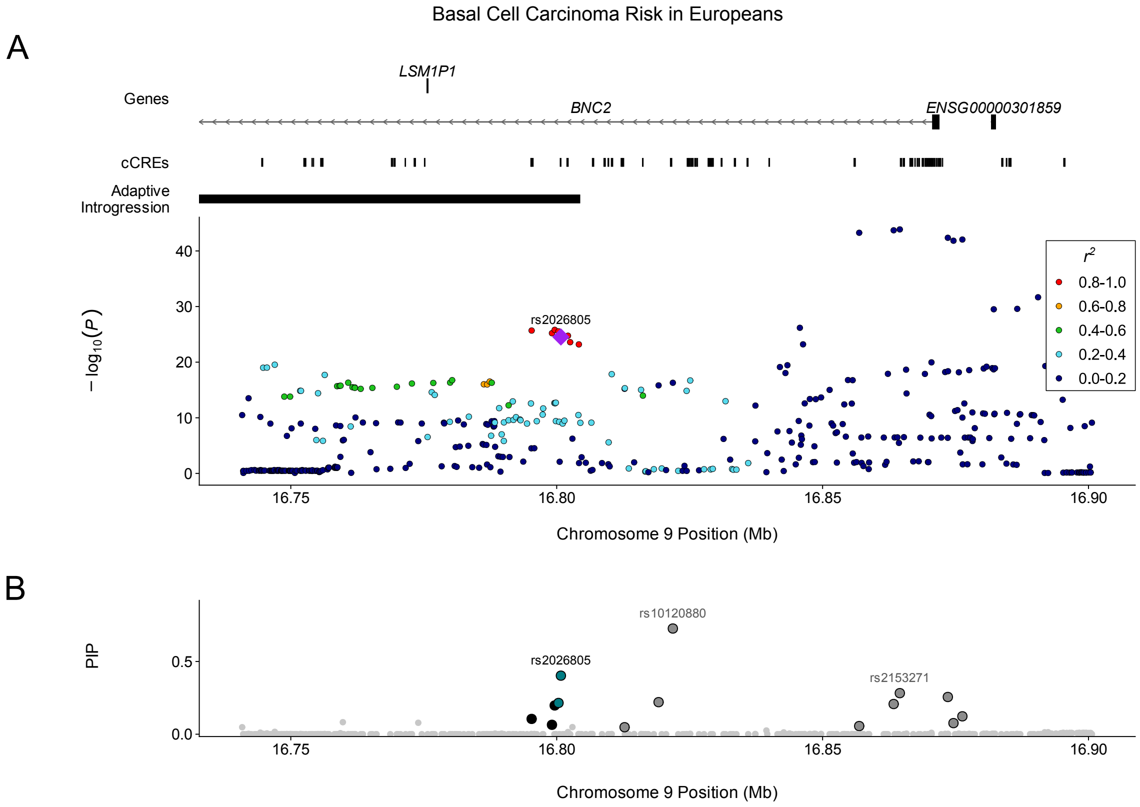


**Fig. S7. Adaptively introgressed Neanderthal variants within *BNC2* associate with increased basal cell carcinoma risk.**
(A) LocusZoom plot of variants from the Seviiri et al. (2022) basal cell carcinoma GWAS conducted among individuals with genetic similarity to Europeans in 1000 Genomes. All variants in the summary statistics within the window are shown. Binned LD ($r^2$) with rs2026805 is indicated by color; variants absent from the 1000 Genomes reference panel, for which $r^2$ could not be calculated, are plotted in the lowest bin. The lead variant, rs2026805, is indicated by a purple diamond. cCREs, as profiled by ENCODE, and adaptively introgressed regions from Iasi et al. (2025) are shown above. (B) Posterior inclusion probabilities (PIPs) for the variants in A. Teal and black mark members of CS2, the credible set this figure concerns, split by whether the variant is Neanderthal (teal) or not (black). Dark grey marks members of two other credible sets at the locus (CS1, CS5) in the shown window, and light grey variants belong to no credible set.

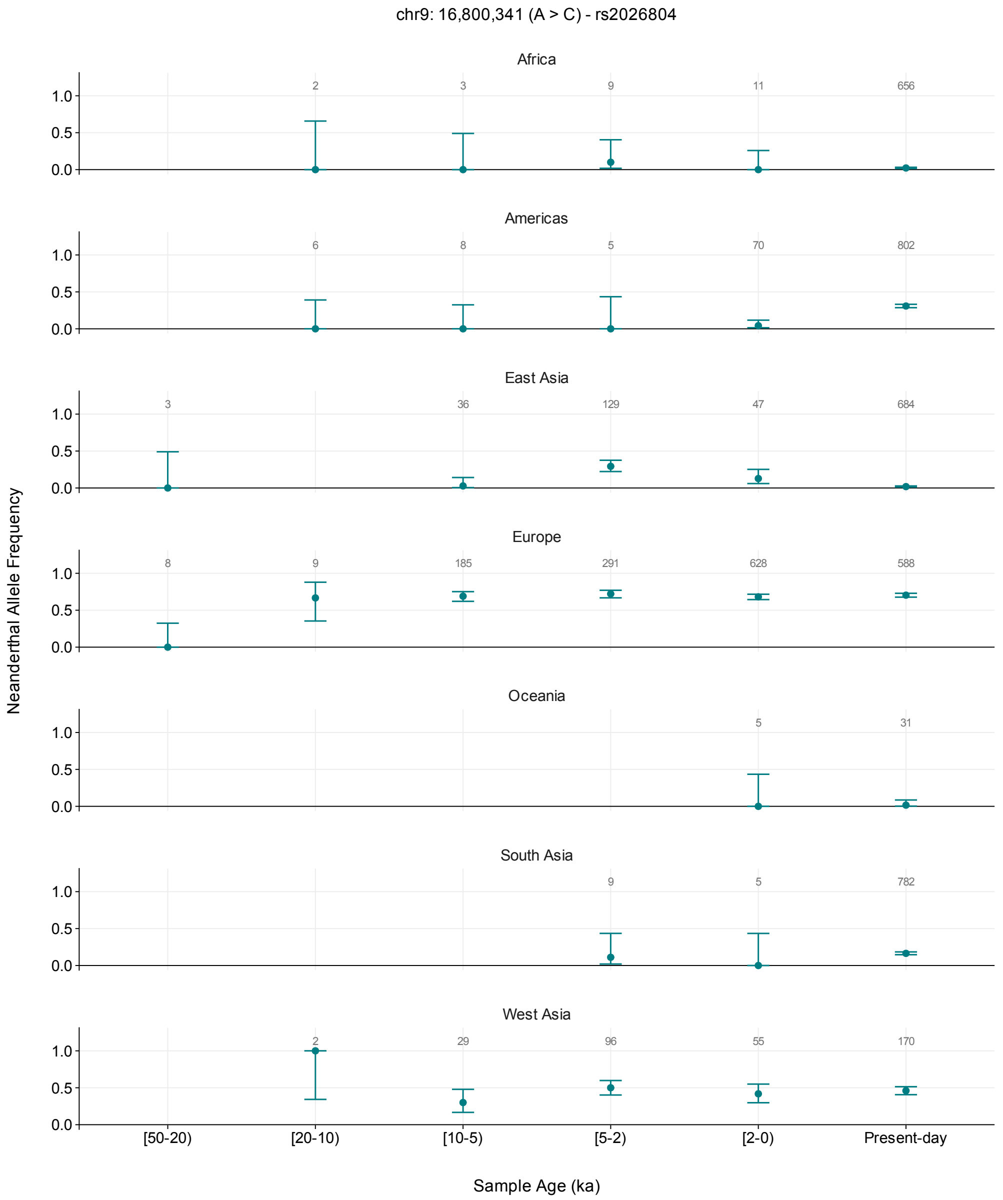


**Fig. S8. rs2026804 is absent from modern human genomes older than 20 ka and is already at high frequency in Europe and West Asia by 10 ka.** Neanderthal allele frequency at rs2026804 (chr9:16,800,341, A>C) in ancient genomes from the Allen Ancient DNA Resource (AADR), binned by sample age and faceted by region. Points give the allele frequency within each bin and error bars the 95% Wilson score interval; the number above each point is the count of specimens contributing to it. Bins are labelled in thousands of years before present and are closed at the older bound and open at the younger, so a specimen dated exactly 2 ka falls in [2-0). One genotype is retained per specimen, preferring diploid shotgun calls, which are the only source able to call a heterozygote; remaining sources are pseudo-haploid and contribute a single allele each. The allele is absent from all 11 modern human specimens older than 20 ka, including 8 from Europe. It is first observed in [20-10), at 6 of 9 alleles in Europe and 2 of 2 in West Asia (Fisher's exact $P = 0.007$ against the preceding bin in Europe); the oldest carrier dates to $\sim$13,700 years ago, and the allele remains near 0.7 in Europe in every subsequent bin. It is absent or rare in Africa, the Americas, Oceania and South Asia in every ancient bin. In East Asia it is near zero in every bin except [5-2), where it reaches 0.293 (39 of 133 alleles from 129 specimens). The carriers in that bin are almost all individuals of West Eurasian Steppe ancestry buried within the region's geographic bounds—Afanasievo, Sintashta, Andronovo, Karasuk and Tagar individuals from southern Siberia, together with Bronze and Iron Age individuals from Xinjiang and western Mongolia—whereas the East Asian-ancestry populations in the same bin carry no copies. AADR regions are geographic rather than genetic labels, so a bin's frequency reflects the ancestry of the populations sampled within it. Present-day samples are shown as a separate category because they are a worldwide anthropological panel rather than a population sample, and their regional frequencies are not comparable to the ancient bins or to continental frequencies from gnomAD or 1000 Genomes.

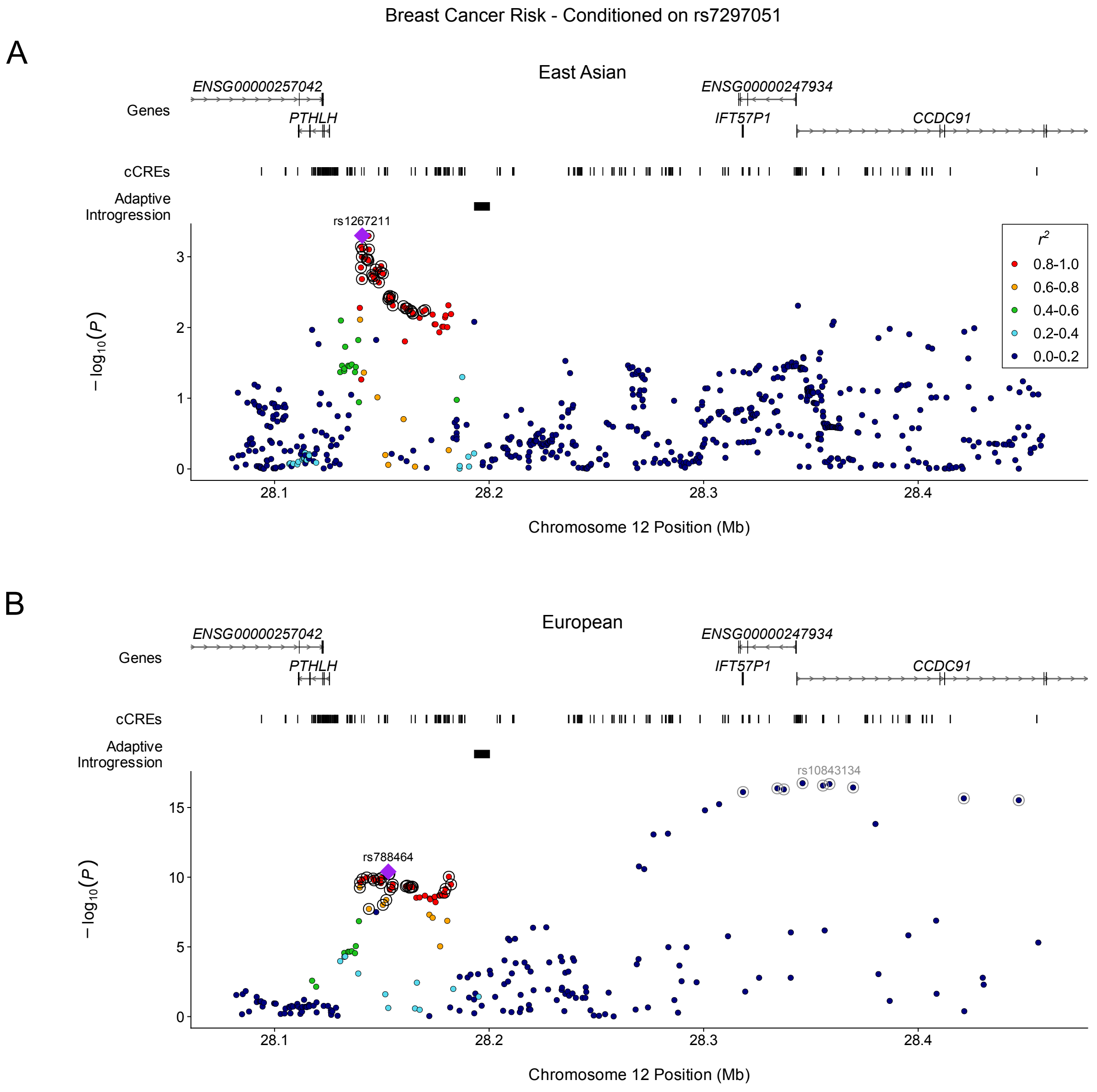


**Fig. S9. A Neanderthal association at *PTHLH* persists after conditioning on the strongest association at the locus in Europeans.**
(A) LocusZoom plot of variants from the Jia et al. (2025) breast cancer GWAS conducted among individuals with genetic similarity to East Asians in 1000 Genomes, after conditioning on rs7297051, computed as $z_{\text{cond}} = (z - rz_{\text{lead}})/\sqrt{1-r^2}$. rs7297051 is the most strongly associated variant at this locus in the European GWAS; the same variant is used in both panels so the two ancestry groups are tested identically. Variants absent from the 1000 Genomes reference panel are excluded, since the conditional statistic requires $r$ with the variant conditioned on. Binned LD ($r^2$) with rs1267211 is indicated by color. rs1267211, the lead variant of the East Asian credible set, is marked by a purple diamond, and the set's remaining members are circled in black. This locus has no other credible set in the East Asian GWAS. Gene models are GENCODE, cCREs are from ENCODE, and adaptively introgressed regions are from Iasi et al. (2025). (B) The same window in the Zhang et al. (2020) breast cancer GWAS conducted among individuals with genetic similarity to Europeans, conditioned on rs7297051 and plotted as in A, with $r^2$ relative to rs788464. rs788464, the lead variant of CS3, is marked by a purple diamond and the set's remaining members are circled in black. Members of the locus's other credible sets are circled in grey, with each set's lead variant labelled in grey: rs7297051, the single member of CS1, and rs10843134, the lead of CS2. The association spanning *PTHLH* persists in Europeans, indicating a signal independent of rs7297051; in East Asians it is attenuated below genome-wide significance (Supplementary Text). CS2 is likewise little affected, its strongest member falling from $|Z| = 8.85$ to 8.51. Both panels span the same coordinates so the two ancestry groups can be compared directly.

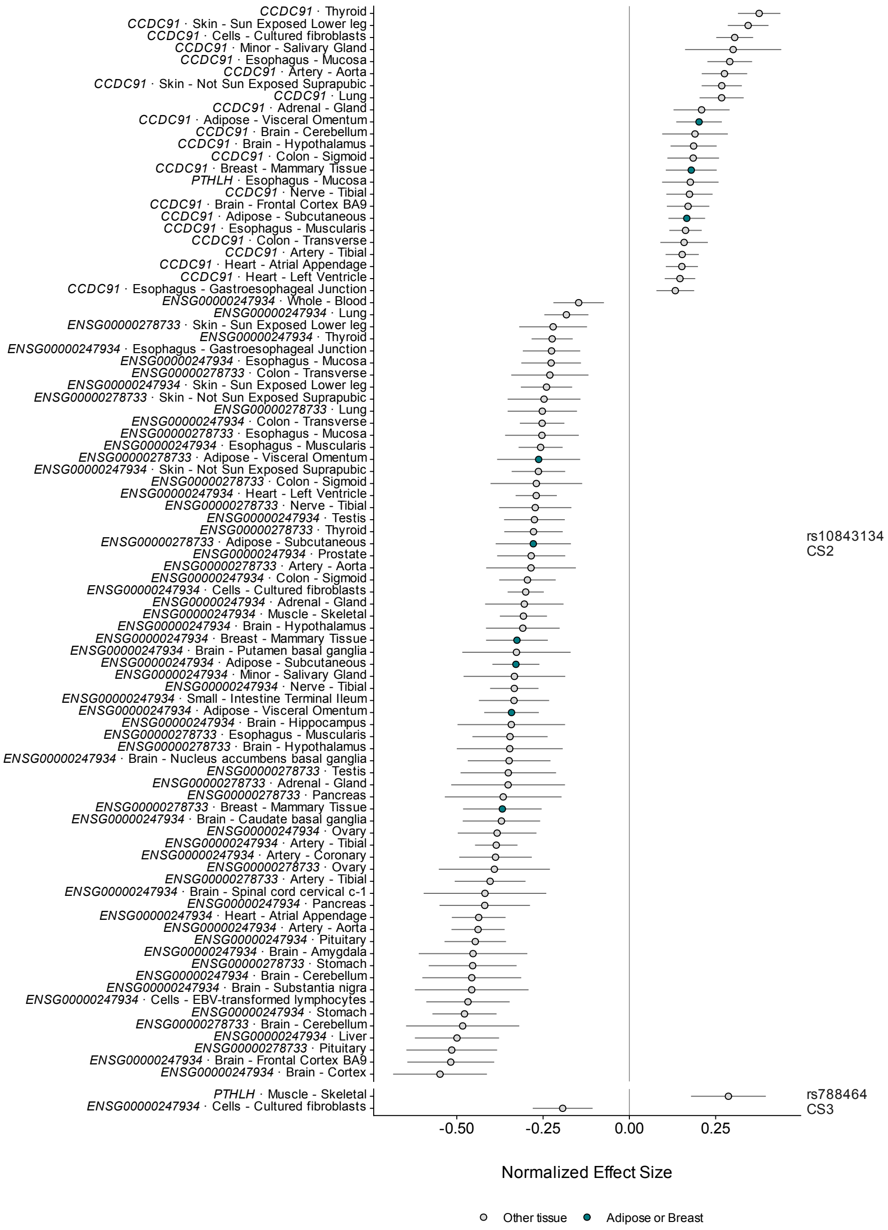


**Fig. S10. Both *PTHLH* breast cancer Neanderthal associations are eQTLs for multiple nearby genes across several tissues.** Normalized effect sizes (NES) for every gene–tissue pair in which rs788464 (CS3) or rs10843134 (CS2) is a significant single-tissue eQTL in GTEx. Points show the NES and lines its 95% confidence interval; effects are oriented to the G allele at both variants, which is the reintroduced ancestral allele at rs788464 and the Neanderthal allele at rs10843134. Adipose and breast tissues are highlighted. Significance is as defined by GTEx.

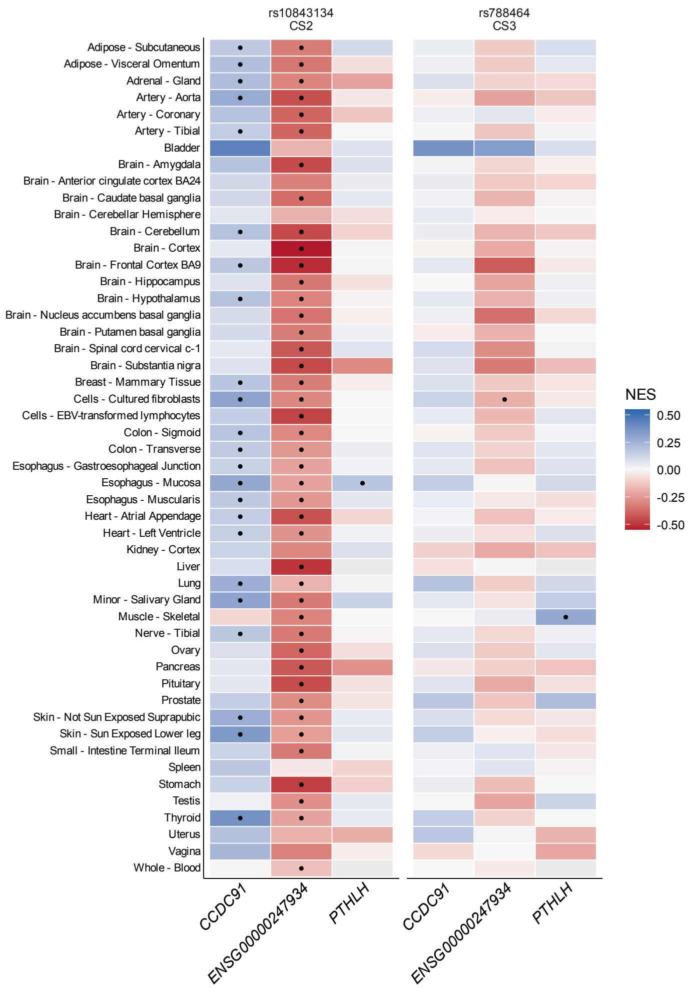


**Fig. S11. Both *PTHLH* Neanderthal associations affect ENSG00000247934 expression.** Normalized effect size (NES) of rs788464 (CS3) and rs10843134 (CS2) on each gene in every GTEx tissue, oriented to the G allele at both variants. Blue indicates that the G allele increases expression and red that it decreases expression. Dots mark significant eQTLs; grey tiles mark variant–gene–tissue combinations that GTEx did not test.

#### Supplementary Tables

Supplementary Tables S1–S17 are provided as separate sheets in a single Excel workbook (`supplementary_tables.xlsx`).

**Table S1.** Candidate cancer GWAS. All 227 East Asian and European cancer GWAS considered, with sample sizes and whether each was selected.

**Table S2.** Cancer GWAS analyzed. The 29 selected GWAS, with sample sizes, effective sample size, genome assembly, numbers of SNVs and genome-wide significant SNVs, and summary statistics URLs.

**Table S3.** Proportion of GWAS loci containing Neanderthal variants, per GWAS, at LD clumping thresholds of $r^2$ = 0.2, 0.5 and 0.8.

**Table S4.** Fine-mapping fit status for each locus.

**Table S5.** Genome-wide significant credible sets from SuSiE-RSS fine-mapping.

**Table S6.** Neanderthal-majority associations: credible sets in which Neanderthal variants carry more than half of the posterior probability.

**Table S7.** Variants in the Neanderthal-majority credible sets, with their Neanderthal allele and its effect size.

**Table S8.** Open Targets credible sets containing *SLC24A4* credible set variants.

**Table S9.** Distance from each Neanderthal-majority credible set to the nearest adaptively introgressed region.

**Table S10.** gnomAD-SV v4 structural variants within the *BNC2* locus (chr9:16,545,287–17,050,791, hg38). Columns report variant ID, chromosome, SV class, position, size, functional consequence, allele count, allele number, allele frequency, and homozygote count. Sorted by allele frequency descending.

**Table S11.** Linkage disequilibrium between structural variants and Neanderthal credible set SNPs at the *BNC2* locus, estimated from SVatalog (1000 Genomes Project long-read data). Columns report SV coordinates (hg19), type, size, allele frequency, the SNP in highest $r^2$ with each SV, and $r^2$ and $D'$ with the lead Neanderthal credible set variant rs2026805.

**Table S12.** Open Targets Genetics variant-to-gene (V2G) scores for Neanderthal credible set variants at the *BNC2* locus.

**Table S13.** gnomAD-SV v4 structural variants within the *PTHLH* locus (chr12:27,736,799–28,696,889, hg38). Columns report variant ID, chromosome, SV class, position, size, functional consequence, allele count, allele number, allele frequency, and homozygote count. Sorted by allele frequency descending.

**Table S14.** Linkage disequilibrium between structural variants and Neanderthal credible set SNPs at the *PTHLH* locus, estimated from SVatalog (1000 Genomes Project long-read data). Columns report SV coordinates (hg19), type, size, allele frequency, the SNP in highest $r^2$ with each SV, and $r^2$ and $D'$ with the lead Neanderthal credible set variant rs7297051.

**Table S15.** Heritability estimation quality control metrics.

**Table S16.** Partitioned heritability enrichment results.

**Table S17.** Kriging diagnostics for fine-mapping model fit.
